# Co-circulation of two viral populations under vaccination

**DOI:** 10.1101/2020.12.29.20248953

**Authors:** Manuel A. Acuña-Zegarra, Mayra Núñez-López, Andreu Comas-García, Mario Santana-Cibrian, Jorge X. Velasco-Hernández

**Affiliations:** Departamento de Matemáticas, Universidad de Sonora, Blvd. Rosales y Luis Encinas S/N, Col. Centro, Sonora, Hermosillo CP 83000, Mexico; Departamento de Matemáticas, ITAM, Río Hondo 1, Ciudad de México CP 01080, Mexico; Departamento de Microbiología, Escuela de Medicina, Universidad Autónoma de San Luis Potosí, Av. Venustiano Carranza 2405, Lomas los Filtros, CP 78210 San Luis, S.L.P; CONACyt - Instituto de Matemáticas UNAM-Juriquilla, Querétaro CP 76230, Mexico; Instituto de Matemáticas UNAM-Juriquilla, Querétaro CP 76230, Mexico

**Keywords:** Mathematical model, Viral populations, Vaccination, Bi-stability, SARS-CoV-2, Influenza

## Abstract

The interaction and possibly interference between viruses infecting a host population is addressed in this work. We model two viral diseases with a similar transmission mechanism and for which a vaccine exists. The vaccine is characterized by its coverage, induced temporary immunity, and efficacy. The population dynamics of both diseases consider infected individuals of each illness and hosts susceptible to one but recovered from the other. We do not incorporate co-infection. Two main transmission factors affecting the effective contact rates are postulated: i) the virus with a higher reproduction number can superinfect the one with a lower reproduction number, and ii) there exists some induced (indirect) protection induced by vaccination against the weaker virus that reduces the probability of infection by the stronger virus. Our results indicate that coexistence of the viruses is possible in the long term, even considering the absence of superinfection. Influenza and SARS-CoV-2 are employed to exemplify this last point, observing that the time-dependent effective contact rate may induce either alternating outbreaks of each disease or synchronous outbreaks. Finally, for a particular parameter range, a backward bifurcation has been observed for dynamics without vaccination.

## 1 Introduction

The interaction between viral species follows known patterns of coexistence. [1] were the first to show that the inclusion of superinfection makes coexistence between competing species possible. Superinfection is a process where a competitive hierarchy exists among a group of species that compete for the same resources. In general (in the absence of vaccines or treatments), this hierarchy facilitates coexistence and prevents competitive exclusion of the weaker species in a process that is mainly driven by the relative magnitude of the reproduction numbers of both species [2] that reflect the abilities to use a contested common resource.

The process of superinfection has been identified as a promoter of the coexistence of pathogen strains in a given host population. The structure of the models that have been used to theoretically demonstrate this property [1, 3, 4], has also been applied to explain the organization of community structure for species that inhabit a common landscape but are not necessarily closely related [5]. For infectious diseases, the concept of superinfection has pervaded theoretical explanations for coexistence, in the same host population, of variants of a given pathogen [6, 7]. The pioneering model in [3] is about infectious diseases and has been used to explore a plausible hypothesis for the length of the latent period of HIV before the onset of AIDS. Simultaneously, these same authors [8] addressed the problem of community structure postulating a trade-off between colonization and extinction [9] in the presence of a hierarchy of competitive abilities for the exploitation of the resources in a common landscape. For acute respiratory infections, this competitive hierarchy that henceforth we will call superinfection has been postulated to explain the alternating dynamics between influenza and RSV (respiratory syncytial virus). It is known that the reproduction number of influenza is higher than that of RSV. This fact, coupled with weather variability (seasonality), induces alternating patterns where influenza and RSV infections have a limited temporal overlap [6]. Seasonal influenza has a median basic reproduction number of 1.28 [10]; while for SARS-CoV-2, the median *R*_0_ is 2.79 [11, 10]. This difference in transmission potential supports the assumption that while competing for hosts, their common resource, the likelihood of long-term coexistence and co-circulation is high and that this balance may be associated with superinfection.

Vaccines have a protective effect via immune responses or indirect effects by reducing the burden of viral and bacterial respiratory diseases on individual patients, among others [12]. However, in general, the impact of vaccination on the transmission of a viral disease starts slowly and builds up over several months to reach target coverage levels. Vaccines are evaluated in terms of efficacy, existing population immunity, coverage and temporary immunity, both natural and vaccine-induced, reduction of mortality or infection risks [13]. Vaccines are applied in order to eradicate a disease. However, this aim depends on many factors and may not be fully achieved.

The present work addresses the general theoretical problem of the co-circulation and long-term persistence or eventual extinction of two viral respiratory infections subject to vaccination. In addition, we explore: i) the order relation between basic reproductive number and vaccine reproduction number, and ii) the existence of backward bifurcation when vaccination is not considered. Finally, we exemplify some of our results with the case of the developing ecological interaction between SARS-CoV-2 and influenza.

The paper is organized as follows. In Section 2 we formulate our mathematical model. In Section 3 we develop the local analysis, including cases of backward bifurcation. In Section 4, we address a particular case, co-infection dynamics between influenza and SARS-Cov-2. Finally, in Section 5 we draw some conclusions about this work.

## 2 Mathematical model set-up

We formulate a mathematical model considering the simultaneous presence of two viruses and vaccination for each one. We do not distinguish between viral subtypes and, therefore, we approximate viral dynamics as if each of them were a single viral population. Both diseases are assumed to present temporary immunity and, thus, the possibility of reinfections. The model is a coupled system of two SIRS (susceptible-infected-recovered-susceptible) equations (Figure 1).

**Figure 1:**
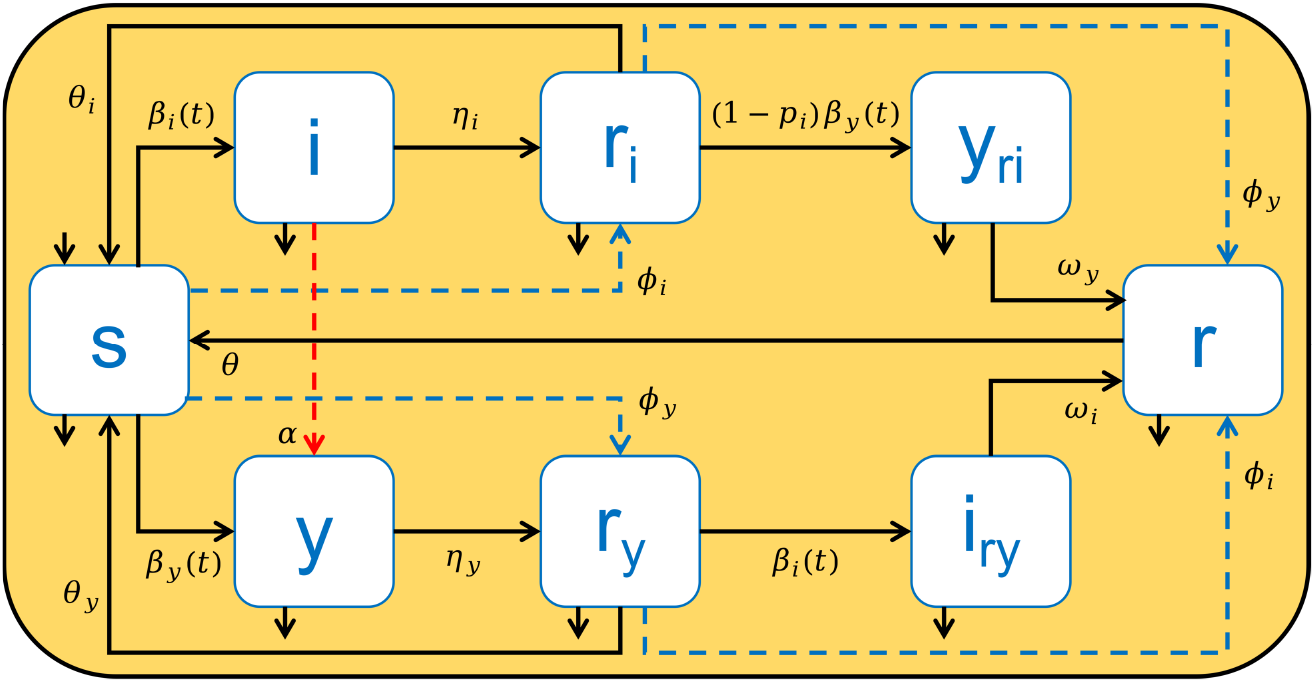
A compartmental mathematical model for two coupled SIRS. 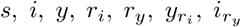 and *r* represent the population of susceptible, infected by virus *i*, infected by virus *y*, immune from virus *i*, immune from virus *y*, immune from virus *i* but infected with virus *y*, immune from virus *y* but infected with virus *i*, and immune from both viruses. Here, *r*_*k*_ includes population recovered after infection and successfully vaccinated by virus *k*, where *k* represents *i* or *y* viruses. Dashed blue lines represent vaccination dynamics for both viruses and dashed red line denotes superinfection process.

We assume a constant total population, normalized to the total population *N*. In the presence of vaccination against both viruses, the equations stand as:

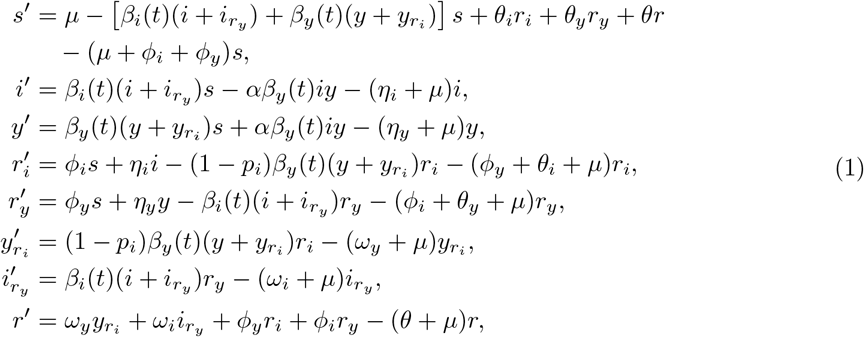

In eq. (1), 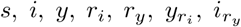 and *r* are defined as described in Figure 1. Note that the immune population for each virus can be infected (vaccinated) by (against) the other virus. With the aim of approximate the seasonal component that drives ILIs [14, 15, 16], for numerical analysis, we assume that the effective contact rates for both viruses, *β*_*i*_(*t*) and *β*_*y*_(*t*), are time-dependent. It is also assumed that virus *y* may potentially competitively exclude virus *i* from its host. The effective contact rate of this transmission route is modulated by *α*, with 0 ≤ *α* ≤ 1 that we define as the superinfection coefficient.

On the other hand, we define *p*_*i*_, 0 ≤ *p*_*i*_ ≤ 1, to measure the indirect protective effects of *i*-vaccinated individual against infection by virus *y* (e.g., as described for influenza and SARS-CoV-2 in [12], additionally [17], found a significant reduction in the odds of testing positive for COVID-19 in patients who received an influenza vaccine compared to those who did not receive the vaccine. In pediatric populations, seasonal influenza and pneumococcal vaccination may have protective value in symptomatic COVID-19 diseases [18]). This protective effect is modelled by a reduction of the effective contact rate of virus *y* when in contact with an immune individual from virus *i* (*r*_*i*_). We assume that there are reinfections by both viruses due to the non-lasting immunity conferred by previous infections. Thus, *θ*_*k*_ is the loss of immunity rate related to virus *k*. To simplify the model, we assume that *θ*, the loss of immunity rate once an individual becomes immune from both viruses, is a constant and does not depend on the order of the infections. Other parameter descriptions are shown in Table 1.

**Table 1:**
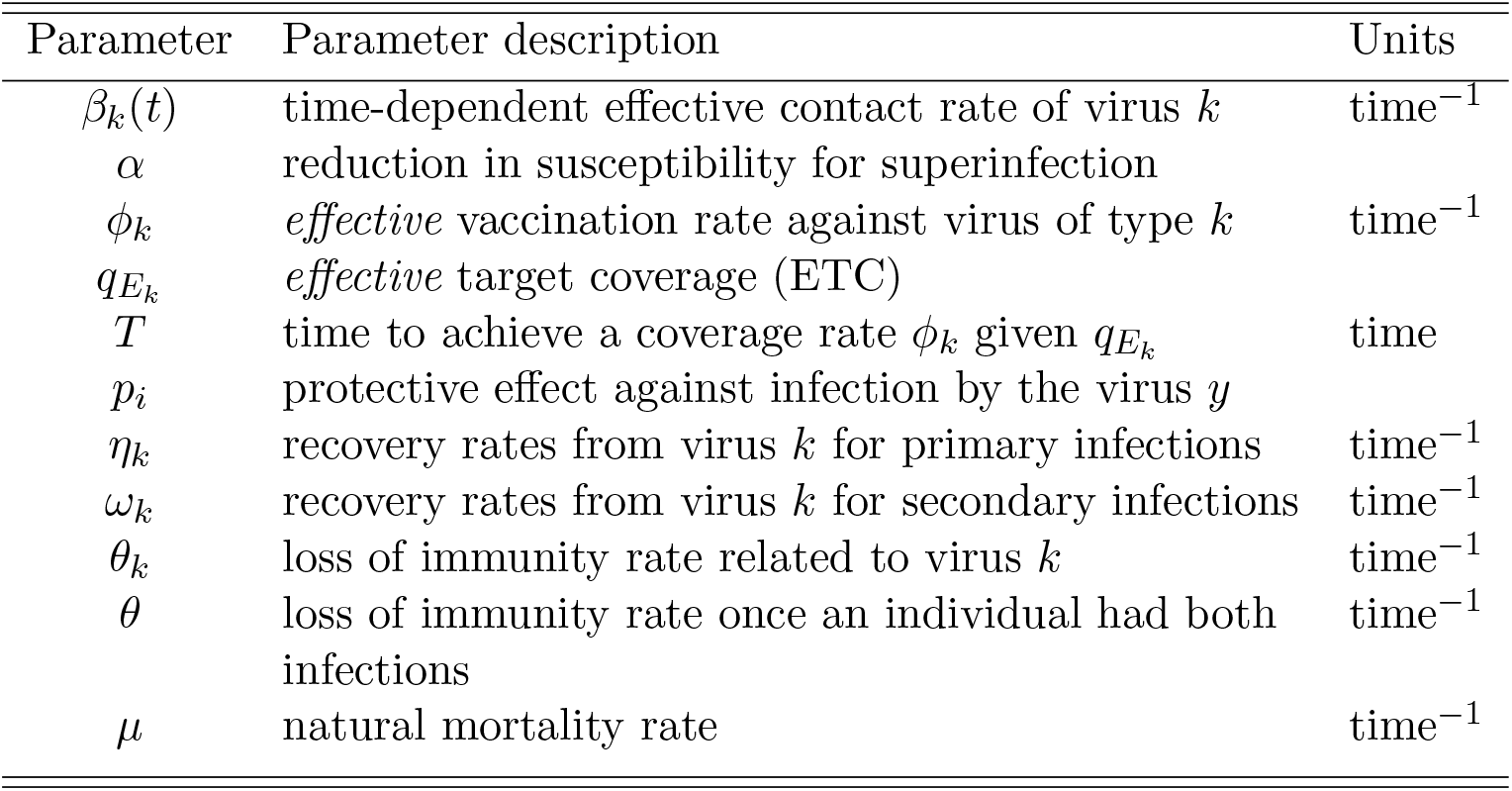
Definition of model parameters for viral populations *k* = *i* and *k* = *y*.

| Parameter | Parameter description | Units |
| --- | --- | --- |
| $\beta_k(t)$ | time-dependent effective contact rate of virus $k$ | $\text{time}^{-1}$ |
| $\alpha$ | reduction in susceptibility for superinfection | |
| $\phi_k$ | *effective* vaccination rate against virus of type $k$ | $\text{time}^{-1}$ |
| $q_{E_k}$ | *effective* target coverage (ETC) | |
| $T$ | time to achieve a coverage rate $\phi_k$ given $q_{E_k}$ | time |
| $p_i$ | protective effect against infection by the virus $y$ | |
| $\eta_k$ | recovery rates from virus $k$ for primary infections | $\text{time}^{-1}$ |
| $\omega_k$ | recovery rates from virus $k$ for secondary infections | $\text{time}^{-1}$ |
| $\theta_k$ | loss of immunity rate related to virus $k$ | $\text{time}^{-1}$ |
| $\theta$ | loss of immunity rate once an individual had both infections | $\text{time}^{-1}$ |
| $\mu$ | natural mortality rate | $\text{time}^{-1}$ |

We do not distinguish between vaccinated and recovered individuals. Instead, we collect immunized individuals into a single compartment; *r*_*i*_ and *r*_*y*_ contain, therefore, those individuals that have either been vaccinated against virus *i* or virus *y*, respectively or that, alternatively, are recovered from a natural infection for either of these two viruses. This modeling choice reduces the system’s dimensionality.

### Effective target coverage definition

We define the vaccination rate, *ϕ*_*k*_ where *k* denotes either virus *i* or virus *y* as an *effective* vaccination rate. In other words, *ϕ*_*k*_ incorporates vaccine efficacy. Under vaccination, susceptible individuals are constantly leaving this compartment at a rate −*ϕ*_*k*_*S*. Thus the probability of having been vaccinated at time *t* is 1 − exp(−*ϕ*_*k*_*t*). Therefore, if we wish that a proportion 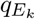 of the susceptible population is vaccinated at time *T*, then we set a vaccination rate such that

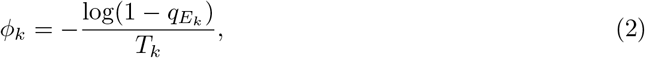

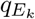 is the *effective target coverage* or ETC with time horizon *T*_*k*_: if the vaccine has *σ*_*k*_% efficacy and we apply the vaccine to *q*_*k*_% of the population, then only a fraction 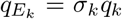 is effectively protected where *k* = *i* denotes virus *i* and *k* = *y* denotes virus *y*.

## 3 Local analysis

First, we briefly characterize some basic properties of the solutions of model eq. (1).

### Lemma 3.1

*Let the initial condition* 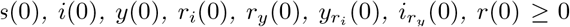. *Then the solution* 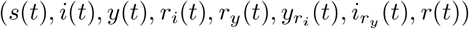 of eq. (1) *is nonnegative for all t >* 0.

The proof is immediate and follows from Proposition A.17 in Appendix A of [19].

### 3.1 Reproduction number

The disease-free equilibrium of eq. (1) always exists and it is given by 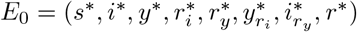 where

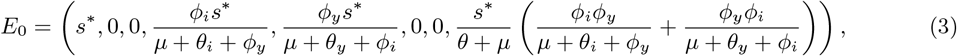

With

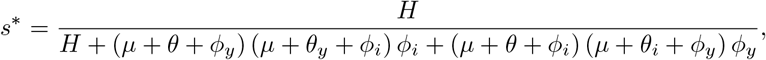

where *H* = (*µ* + *θ*) (*µ* + *θ*_*i*_ + *ϕ* _*y*_) (*µ* + *θ*_*y*_ + *ϕ* _*i*_). Note that the susceptible population at the disease-free equilibrium depends on the parameters for coverage and immunity for both viruses.

Given the interaction of both viruses, their reproduction numbers give information on conditions for coexistence, competitive exclusion, or extinction. In what follows, we give a first characterization for these properties. When the diseases have not yet invaded the host population, but the host is vaccinated against both viruses, we compute the vaccine reproduction number. We proceed as in [20] to obtain:

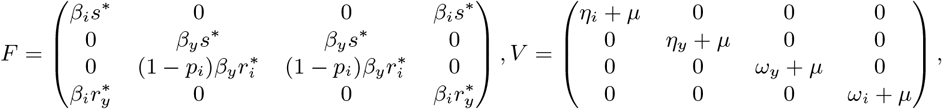

Here, 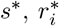 and 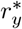 are defined in eq. (3). Then, the vaccine reproduction number is given by the spectral radius of matrix *FV* ^−1^:

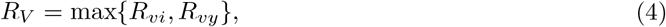

where the vaccine reproduction numbers for virus *i* (*R*_*νi*_) and virus *y* (*R*_*νy*_) are:

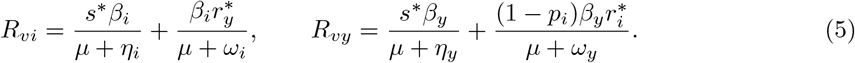

**Remark:** In the absence of vaccination for either one of the viruses, and for constant effective contact rates, the basic reproductive numbers are the classical expressions of an SIR epidemic, that is

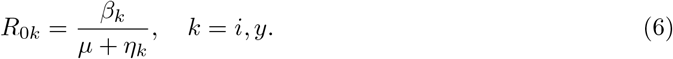

From eqs. (5) and (6), the order relation kept by *R*_*νk*_ and *R*_0*k*_, with *k* = *i, y*, is not obvious. For example, Figure 2 shows that taking *µ* = 0.000039139, *β*_*i*_ = 0.3, *β*_*y*_ = 0.2, *η*_*i*_ = 1*/*5, *η*_*y*_ = 1*/*14, *θ* = 1*/*365, *θ*_*i*_ = 1*/*365, *θ*_*y*_ = 1*/*180, *ω*_*i*_ = 1*/*7, *ω*_*y*_ = 1*/*16, *p*_*i*_ = 0.05. Varying both coverages 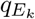 from 1% to 99% (eq. (2)), it is possible that *R*_*νk*_ *> R*_0*k*_ with *k* = *i* or *y*, i.e, vaccine application may enhance rather than reduce the occurrence of an outbreak. It is therefore, important to find the conditions that guarantee that only a reduction occurs, i.e., that *R*_*νk*_ *< R*_0*k*_. In Figure 2, the time horizon to reach vaccination coverages against viruses *i* and *y* is fixed on four and three months, respectively.

**Figure 2:**
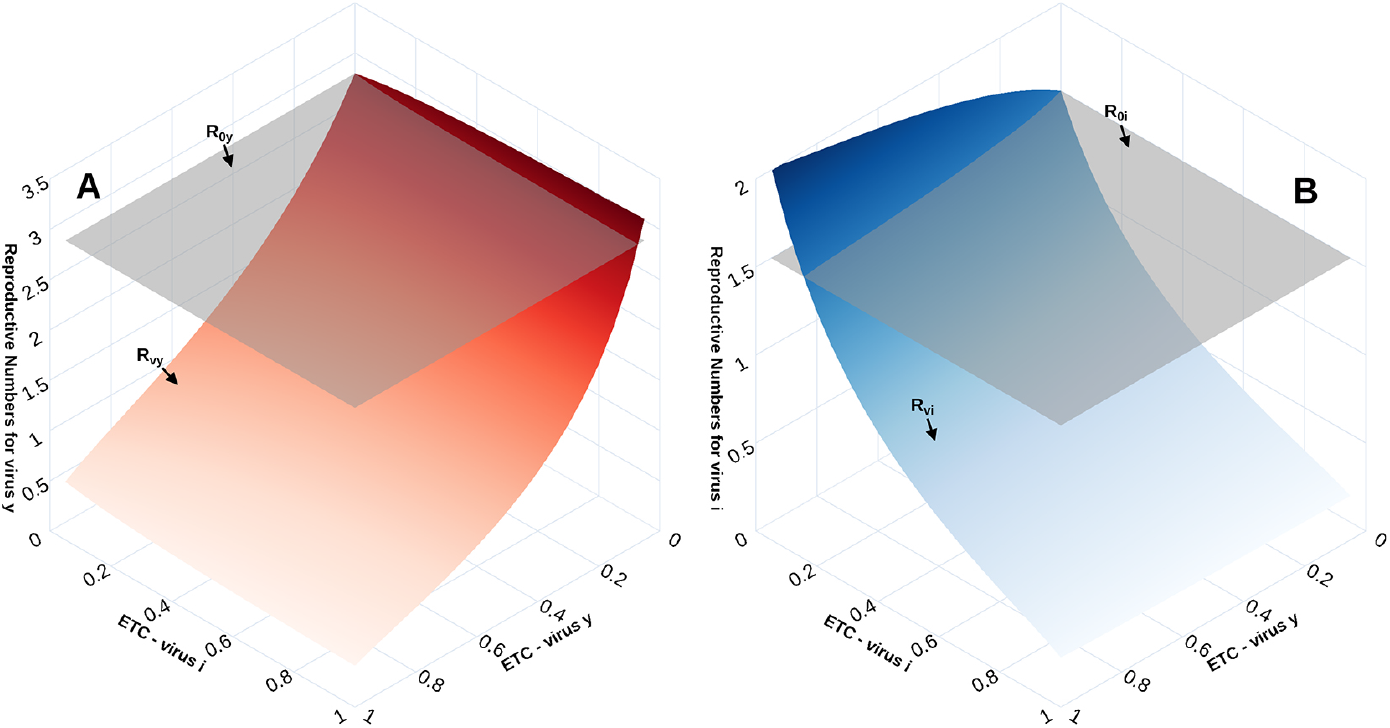
Basic reproduction number vs vaccine reproduction number. Panel A – virus *y*. Panel B -virus *i*. In both cases, planes represent the basic reproduction number (6). Vaccine reproduction numbers (5) are plotted as functions of the effective target coverage (ETC).

We further characterize the relationship between these reproductive numbers.

#### Lemma 3.2

*Let R*_*νk*_ *and R*_0*k*_ be defined as in eqs. (5) and (6), *respectively; and* 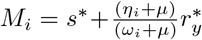 *and* 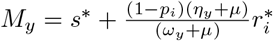

1. min{*M*_*i*_, *M*_*y*_} *>* 1 *if and only if R*_*νk*_ *> R*_0*k*_, *with k* = *i, y*.
2. max{*M*_*i*_, *M*_*y*_} *<* 1 *if and only if R*_0*k*_ *> R*_*νk*_, *with k* = *i, y*.

*M*_*i*_ and *M*_*y*_ can be interpreted as the effective proportion of susceptible individuals available to infection by virus *i* and *y*, respectively. This lemma underlines the dependence of the order relation between the reproductive numbers *R*_*νk*_ and *R*_0*k*_, with *k* = *i, y*, on the pool of susceptible individuals for each virus.

#### Lemma 3.3

*The disease-free equilibrium E*_0_ (eq. (3)*) is locally asymptotically stable if and only if R*_*ν*_ *<* 1.

The proof is given in Appendix A.

### 3.2 When *R*_0*k*_ *< R*_*νk*_

In lemma 3.2, we have shown that there is a range of parameter combinations such that *R*_0*k*_ *< R*_*νk*_, with *k* = *i, y*, that is, where vaccination is not effective in reducing transmission. In this section we explore this case. We fix *µ* = 0.000039139, *α* = 0.5, *η*_*i*_ = 1*/*5, *η*_*y*_ = 1*/*14, *θ* = 1*/*365, *θ*_*i*_ = 1*/*365, *ω*_*i*_ = 1*/*21, *ω*_*y*_ = 1*/*56, *p*_*i*_ = 0, *ϕ* _*i*_ = 0.001603, *ϕ* _*y*_ = 0.002341 and *θ*_*y*_ = 1*/*180. Thus, *M*_*i*_ = 1.131837 and *M*_*y*_ = 1.076793.

Figure 3 shows the asymptotic equilibria, when *t* = 36500, of *i* and *y* as function of *R*_*νi*_ and *R*_*νy*_. Both effective contact rates values are constant and defined such that 0.4 ≤ *R*_*νi*_ ≤ 1.6 and 0.5 ≤ *R*_*νy*_ ≤ 2, with the initial condition (*s*(0), *i*(0), *y*(0), *r*_*i*_(0), *r*_*y*_(0), *i*_*ry*_(0), *y*_*ri*_(0), *r*(0)) = (0.8498, 0.0001, 0.0001, 0.1, 0.05, 0, 0, 0). Figure 3 illustrates that when *R*_*νk*_ *>* 1, with *k* = *i, y*, both disease coexist. Moreover, there exist combinations of parameter values such that *R*_*νi*_ *<* 1 *< R*_*νy*_ which also implies coexistence (Figure 3A). This phenomenon can be interpreted as a kind of rescue effect of one virus by the other. Figure 3B also shows this phenomenon. Finally, when *R*_*νk*_ *<* 1, with *k* = *i, y*, both disease go extinct.

**Figure 3:**
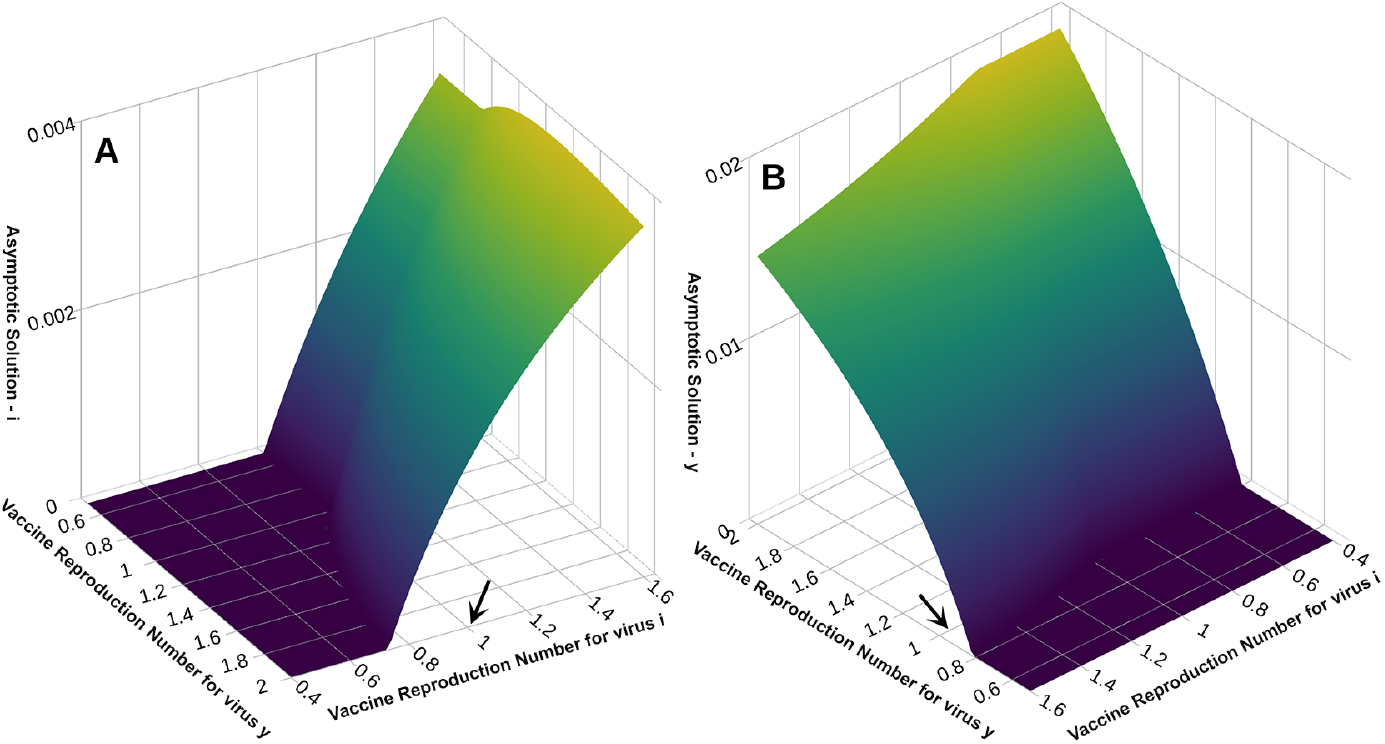
Asymptotic equilibria for *t* = 36500, of *i* and *y* as functions of *R*_*νi*_ and *R*_*νy*_. Panel A: asymptotic prevalence of *i*. Panel B, asymptotic prevalence of virus *y*. In both panels, the black arrow points to the threshold value of 1. There exist parameters values allowing coexistence even if *R*_*νk*_ *<* 1, with *k* = *i* or *y*.

Figure 4 shows a viral coexistence. Here, *β*_*i*_ and *β*_*y*_ are 0.19 and 0.07, respectively. This case shows that even when both basic reproductive numbers *R*_0*k*_ are less than one, both viruses persist in the absence of vaccination. This pattern strongly indicates the existence of bi-stability and of a backward bifurcation [21]. Figure 5 confirms the existence of bi-stability. Here, we fix initial conditions (*s*(0), *i*(0), *y*(0), *r*_*i*_(0), *r*_*y*_(0), *i*_*ry*_(0), *y*_*ri*_(0), *r*(0)) = (0.8499 − *y*_0_, 0.0001, *y*_0_, 0.1, 0.05, 0, 0, 0). When *y*_0_ = 0.0003 both viruses become extinct and the corresponding eigenvalues are (−0.04765819, −0.01789628, −0.01003914, −0.00559469, −0.00277887, −0.00277887, −0.00146771, −0.00003914). Coexistence is observed when *y*_0_ = 0.00032. For this behavior, the eigenvalues are (−0.1369964, −0.04611382, −0.01838541, −0.00176767 + 0.01417291*i*, −0.00176767 −0.01417291*i*, −0.00038391 +0.00493473*i*, 0−.00038391 0−.00493473*i*, 0− .00003914).

**Figure 4:**
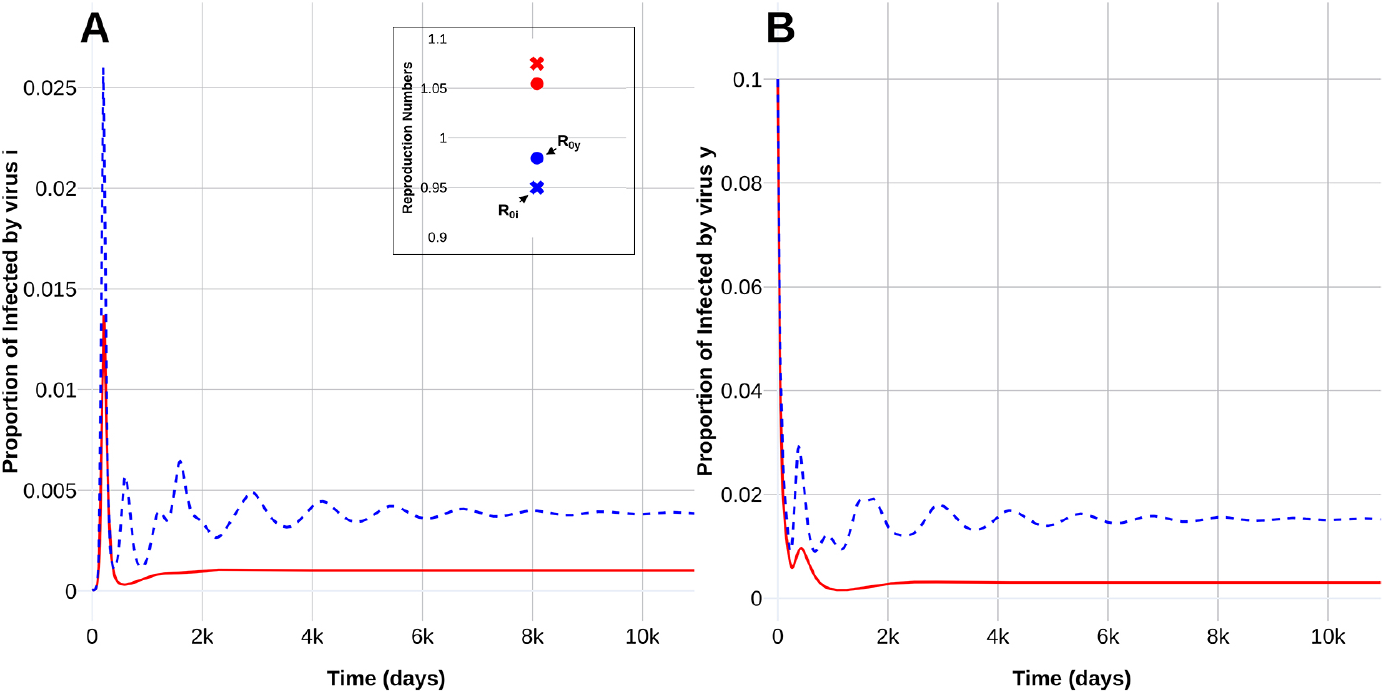
Coexistence of viruses *i* and *y* when 1 *< R*_*νi*_, *R*_*νy*_. A) Virus *i*, B) Virus *y*. Solid red lines: with vaccination. Dotted blue lines, no vaccination. Coexistence occurs even in the absence of vaccination. In the inset dots represent reproduction numbers for virus *y*, and crosses represent reproduction numbers for virus *i*. Color codes: Red vaccination is applied, blue, no vaccination.

**Figure 5:**
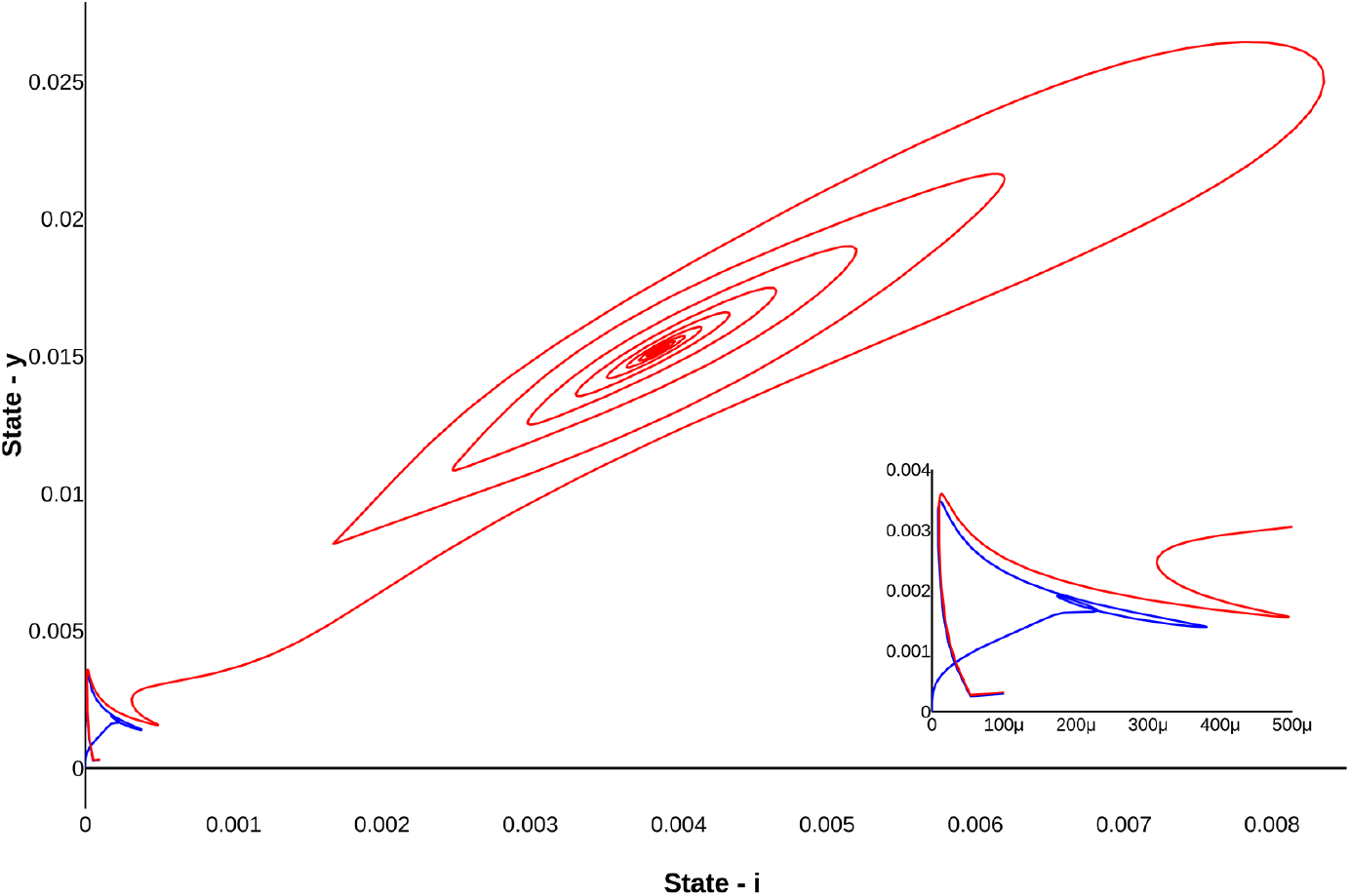
Bi-stability when *R*_0*k*_ *<* 1, with *k* = *i, y*. Red line represents coexistence of both viruses. Blue line shows extinction. The inset is a zoom of the extinction dynamics.

A backward bifurcation means that the reproduction number being less than unity becomes only a necessary, but not sufficient condition, for disease elimination. To further explore the existence of the backward bifurcation shown in Figures 4 and 5, we performed the numerical continuation of the equilibrium points in appropriate parameter regions.

Figure 6 shows the numerical continuation of the equilibrium points of coexistence and extinction for both viruses without vaccination. The continuation is carried out by varying the values of *β*_*k*_ so that *R*_0*k*_ is below and above unity. All numerical results were implemented in Matcom (Matlab tool) [22].

**Figure 6:**
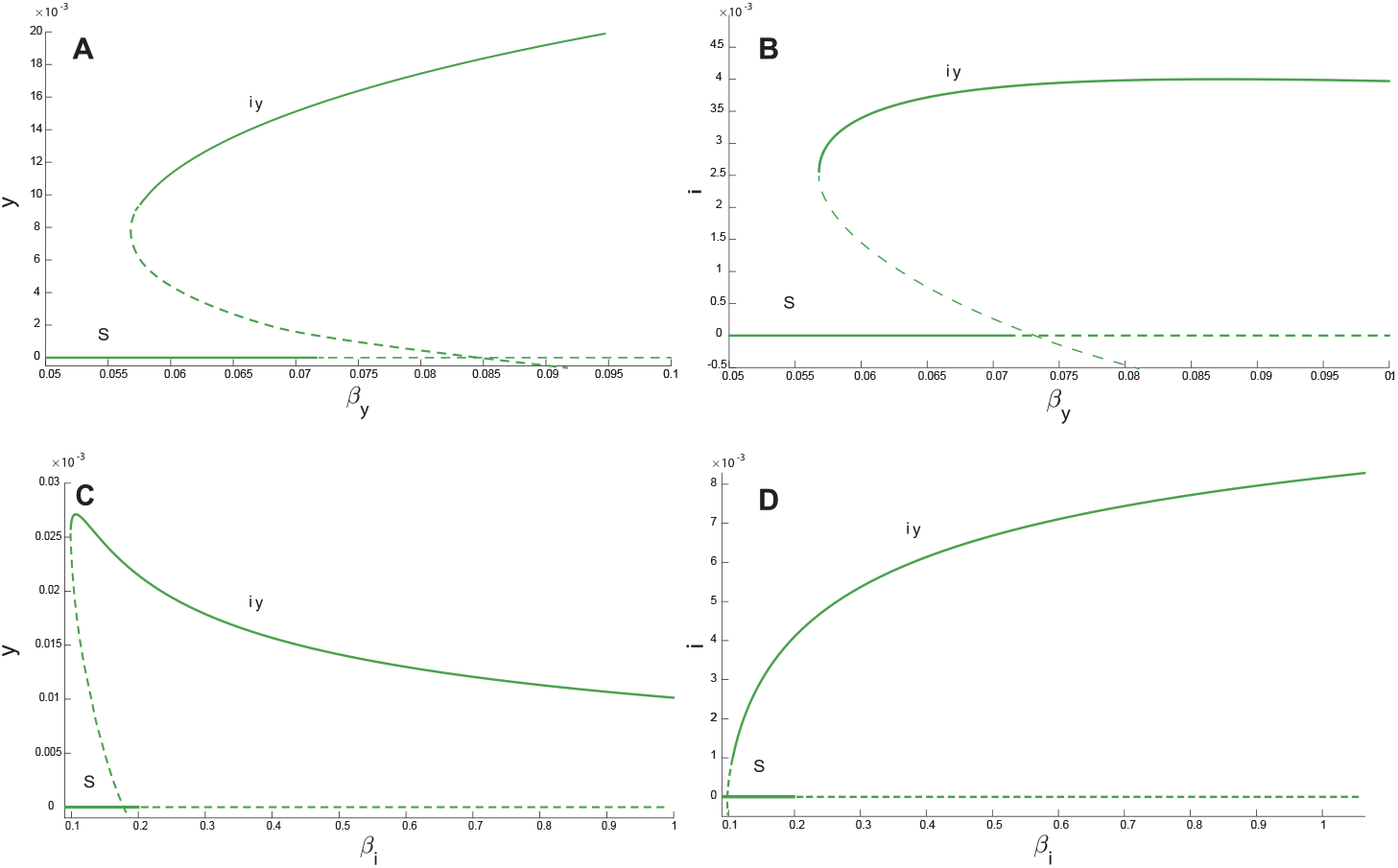
Numerical continuation of equilibrium points for viral coexistence and extinction of both viruses without vaccination as a function of effective contact rate *β*_*k*_ and virus *k. s* represents the continuation extinction curve of both viruses and *iy* the coexistence curve. A) *s* and *iy* projected onto the plane *β*_*y*_ and *y*. B) Plane *β*_*y*_ and *i*. C) Plane *β*_*i*_ and *y*. D) Plane *β*_*i*_ and *i*.

Figure 6 illustrates the numerical continuation of equilibrium points for viral coexistence and extinction of both viruses (in the absence of vaccination), projected on the plane *β*_*k*_ and virus *k* for *k* = *i, y*. Parameter values are *β*_*i*_ = 0.19, *α* = 0.5, *θ*_*i*_ = 1*/*365, *θ*_*y*_ = 1*/*180, *θ* = 1*/*365, *η*_*i*_ = 1*/*5, *η*_*y*_ = 1*/*14, *ω*_*i*_ = 1*/*21, *ω*_*y*_ = 1*/*56, *p*_*i*_ = 0. Figure 6A shows that the continuation extinction curve of both viruses given by *s* is locally stable (solid line) up to the value *β*_*y*_ = 0.0714676, corresponding to a reproduction number of *R*_0*y*_ = 1.0054. In curve *s* as the virus *y* infection rate increases, the disease-free equilibrium curve becomes unstable (dashed line). The coexistence curve of both viruses denoted by *iy*, is locally stable (solid line) for *β*_*y*_ ∈ [0.05814, 0.2697] or, equivalently for 0.81 *< R*_0*y*_ *<* 3.77.

In Figure 6B, we show the numerical continuation of equilibrium points: for viral coexistence and extinction of both virus projected on the plane *i* − *β*_*y*_. The disease-free equilibrium branch *s*, is locally stable (solid line) until *R*_0*y*_ = 1.0054. The stability interval of the branch with the two virus present, *iy* (solid line) is 0.81 *< R*_0*y*_ *<* 3.79. Figs. 6C-D illustrate the numerical continuation of equilibrium points (coexistence and extinction of both virus) projected on the planes *y* − *β*_*i*_ and *i* − *β*_*i*_, respectively. In both, the disease-free equilibrium branch *s* is locally stable (solid line) until *R*_0*i*_ = 1.09, the stability of the coexistence curve starts at *R*_0*i*_ = 0.502.

In all cases, we show that a stable coexistence equilibrium exists together with a disease free equilibrium when *R*_0*k*_ *<* 1. The usual causes of backward bifurcation in some standard deterministic models are imperfect vaccination [23, 24], the existence of exogenous re-infections [25] or vaccine-derived immunity waning at a slower rate than natural immunity [26], the role of re-infection [27], among others [28]. Since the recovery rate from the second infection for both viruses is smaller than the recovery rate from the first infection, the influx to the pool of susceptibles occurs in a time window larger than expected if only one infection were present. This continuous and extended influx generates a backward bifurcation. Therefore the vaccination strategy must be efficient (large coverage in as short a time as possible), to prevent the increase in the pool of susceptibles that may lead to undesirable outcomes.

In general, as superinfection increases in strength (*α* increases), we observe a corresponding slight decrease in the number of superinfected (with virus *i*) hosts and an increase in the superinfector (with virus *y*). This is consistent with standard results, [1, 3, 4]. However, there exist parameter values where an increase of *α* produces an unexpected change in disease dynamics.

Figure 7 shows the effect of increasing superinfection (*α*) for a particular initial conditions and parameter values. As an example we take (*s*(0), *i*(0), *y*(0), *r*_*i*_(0), *r*_*y*_(0), *i*_*ry*_(0), *y*_*ri*_(0), *r*(0)) = (0.8496, 0.0001, 0.0003, 0.1, 0.05, 0, 0, 0). Other parameters are as in Figure 5 to insure that *R*_0*k*_ *<* 1, with *k* = *i, y*. Apparently, there is a threshold value for *α* that switches the dynamics from coexistence to extinction in both viruses. For example, if we consider *α* = 0.5 the corresponding eigenvalues are (−0.04765819, −0.01789628, −0.01003914, −0.00559469, −0.00277887, −0.00277887, −0.00146771, −0.00003914) which is consistent with the behavior shown in Figure 7. On the other hand, when *α* = 0.4, we observed coexistence and the corresponding eigenvalues are (−0.1369048, −0.04614704, −0.01837886, −0.00176734 + 0.01417675*i*, −0.00176734 − 0.01417675*i*, −0.00038428 + 0.00493486*i*, −0.00038428 − 0.00493486*i*, −0.00003914).

**Figure 7:**
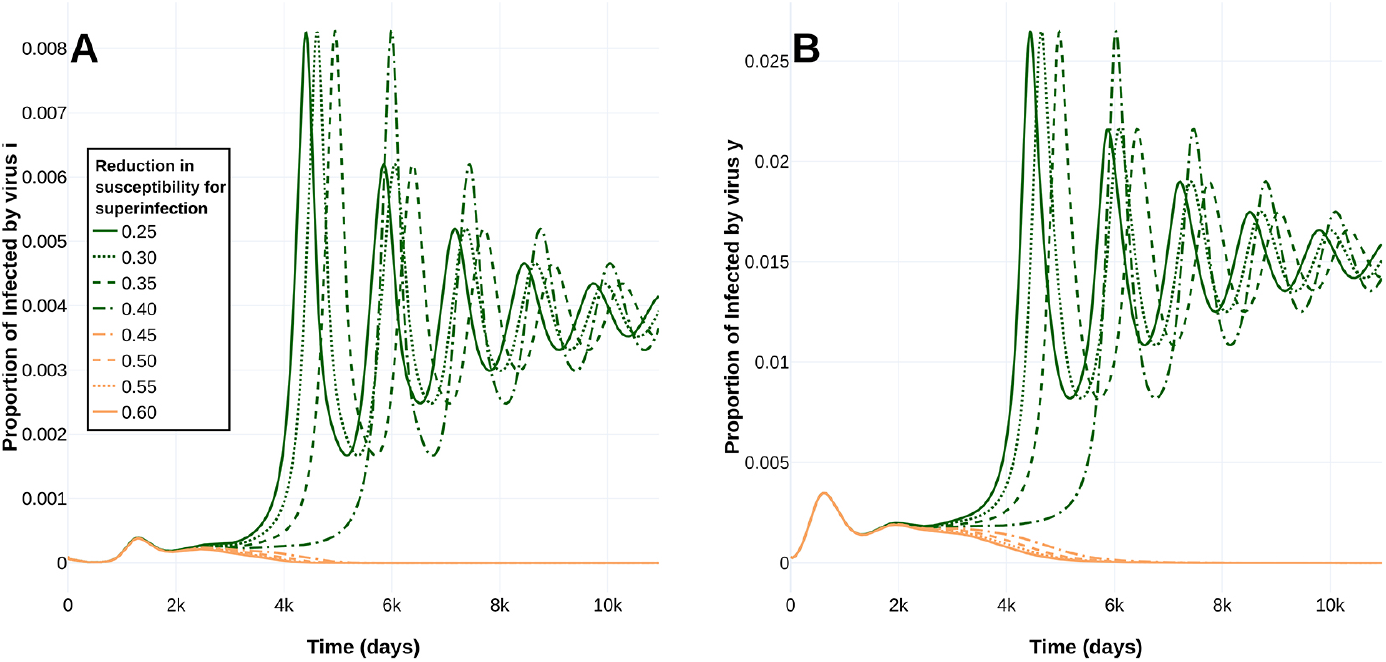
Effect of increasing the superinfection index (*α*) when *R*_0*k*_ *<* 1, with *k* = *i, y*. A) Virus *i*, B) Virus *y*. Extinction occurs when *α* = 0.45, 0.50, 0.55 and 0.60. Coexistence occurs for *α* = 0.25, 0.30, 0.35 and 0.40. *s* represents the extinction curve of both viruses and *xy* the coexistence curve.

## 4 A particular case: Influenza and SARS-CoV-2

The year 2020 and early 2021 have been atypical, at least in two ways. One is the SARS-CoV-2 pandemic becoming the dominant and most prevalent respiratory viral infection from the beginning of 2020. The other is the characteristic absence of a significant number of influenza cases; influenza activity has been almost null in the southern hemisphere and now in the northern, while the SARS-CoV-2 pandemic is active [29]. Figure 8 exemplifies this in the case of Mexico. The winter months (Northern hemisphere) did not produce influenza outbreaks concurrent with COVID-19 resurgence and, thus, hospital capacity in Mexico and elsewhere was not compromised [30, 31]. A possible explanation for the absence of influenza is that the measures are taken to prevent COVID-19 (social distancing, mask use, etc.) also prevent influenza transmission. These measures have limited ability to stop the COVID-19 (aerosols) but may be quite effective at preventing influenza transmission. Nevertheless, the occurrence of a syndemic episode with co-circulating influenza and SARS-CoV-2 viruses is still a potential reality.

**Figure 8:**
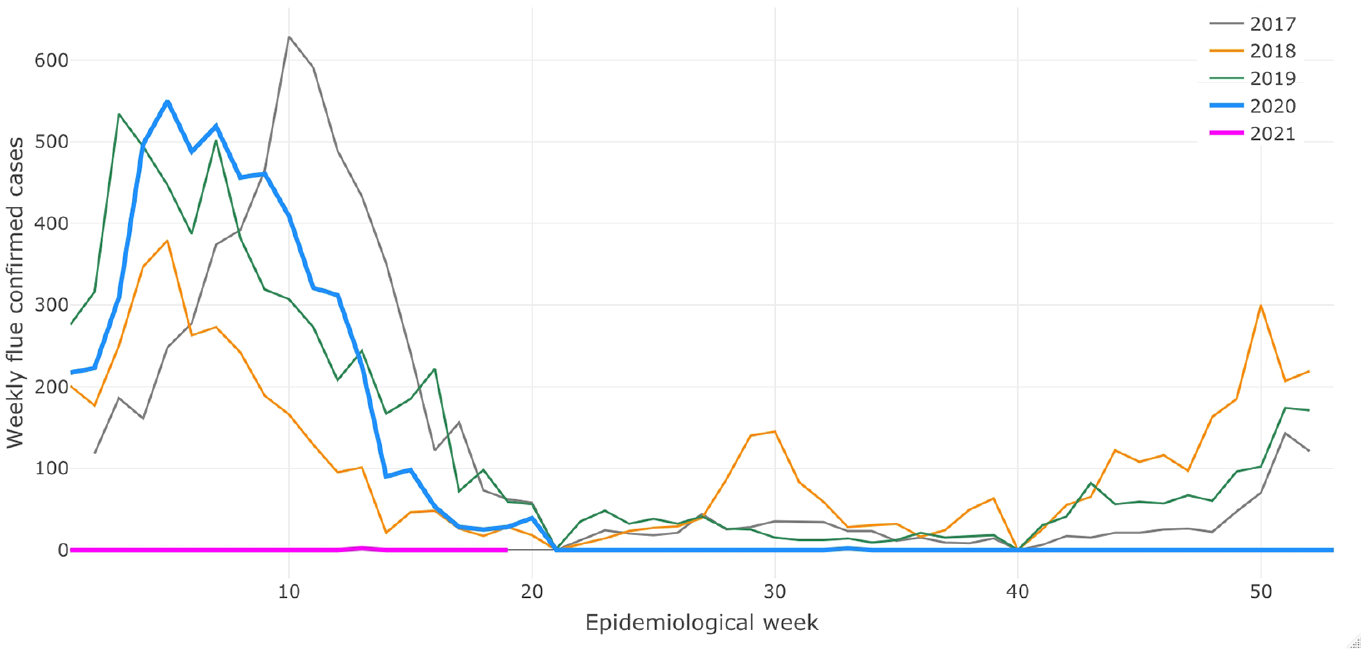
Atypical influenza evolution. Number of weekly influenza confirmed cases by epidemiologcal week from 2017 to 2021. Notice that the number of cases since the 22nd epidemiological week (late May 2020) is almost zero, which is atypical compared to previous years.

In this section, Eq. (1) is used to explore co-circulation dynamics between SARS-CoV-2 and influenza. Given that, to date, there exist no evidence to suggest that a COVID-19 infection has been observed to displace an influenza infection, we consider *α* = 0. Baseline parameters are given in Appendix B.1.

### 4.1 Reproduction numbers, vaccine efficacy, effective coverage and temporary immunity

The dependence of the vaccine reproduction number (eq. (4)) on vaccination coverage and temporary immunity is explored now. For the rest of this section, we fix *µ* = 0.000039139, *η*_*i*_ = 1*/*5, *η*_*y*_ = 1*/*14, *θ* = 1*/*365, *θ*_*i*_ = 1*/*365, *ω*_*i*_ = 1*/*5 and *ω*_*y*_ = 1*/*14. The ETC for vaccine 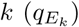 determines the corresponding *ϕ* _*k*_ through eq. (2), with *k* = *i, y*. We fix the effective transmission rates to *β*_*i*_ = 0.32 and *β*_*y*_ = 0.15.

Figure 9 shows *R*_*V*_ as function of ETC for SARS-CoV-2 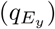 achieved in three months (see eq. (2)) and the average duration of immunity by SARS-CoV-2 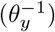. The protective effect of the influenza vaccine (*p*_*i*_) are also left free to vary. As expected, the maximum value of *R*_*ν*_ is achieved when there is not vaccination, that is, 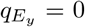. Likewise, we also observe that as increases *p*_*i*_ the maximum value of *R*_*v*_ is lower.

**Figure 9:**
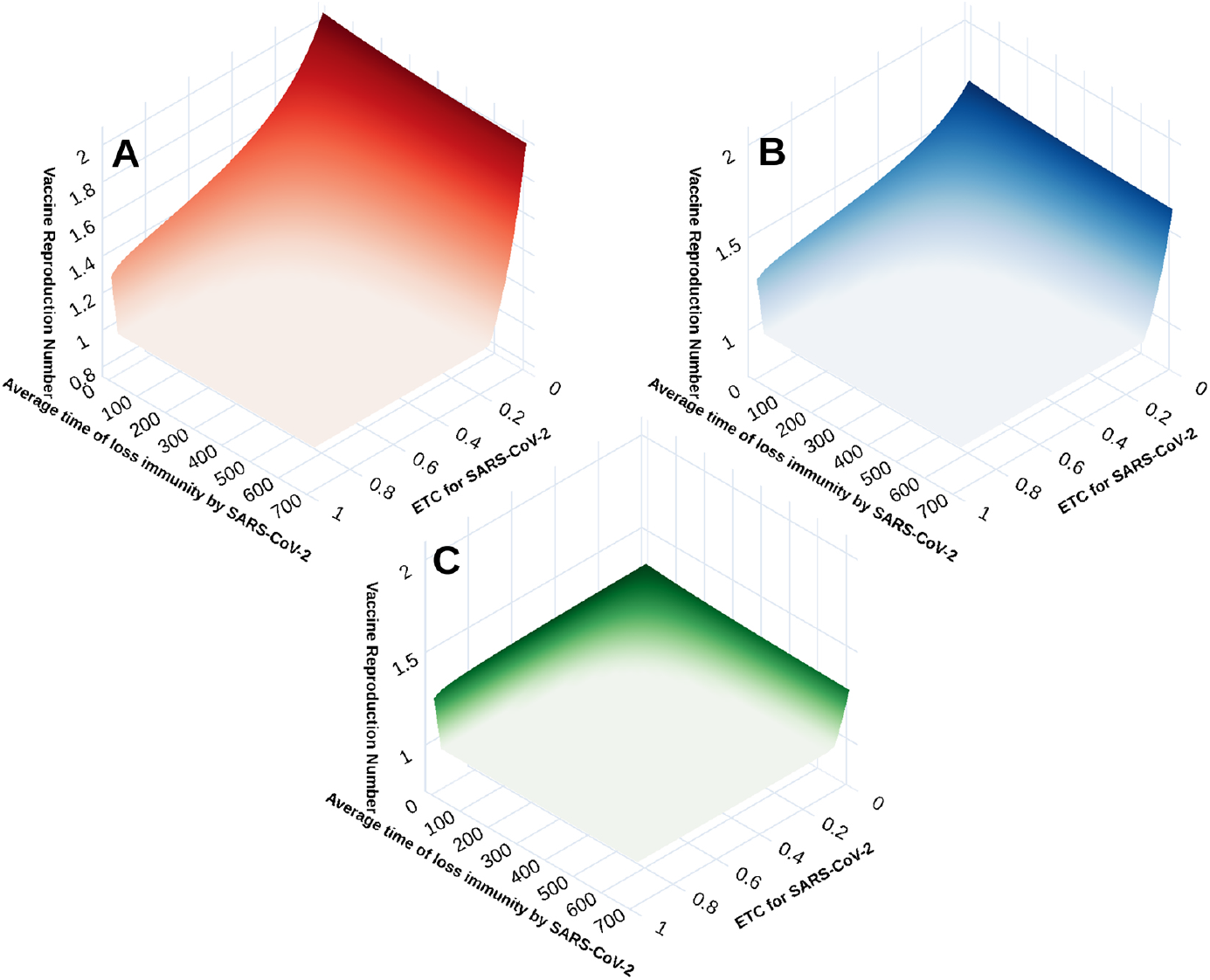
Vaccine reproduction number as a function of coverage and average duration of immunity for SARS-CoV-2. A) No protection from influenza vaccination (*p*_*i*_ = 0). B) 50% protection (*p*_*i*_ = 0.5). C) Full protection (*p*_*i*_ = 1). Target coverage (TC) for influenza is 35% in 4 months with a vaccine efficacy of 50%. Note that the greater *p*_*i*_ is, the higher the reduction in *R*_*V*_.

The effect of the vaccine efficacy and time horizon over vaccine reproduction numbers are presented in Appendix B.2.

### 4.2 Transmission reduction by vaccination (*R*_*νk*_ *< R*_0*k*_)

Numerical explorations of eq. (1) allows us to postulate the diagram in Figure 10. We observe that when both vaccine reproductive numbers are less than one, both epidemics die out. When only one of the vaccine reproductive numbers is greater than one, then the virus associated with that reproductive number persists and the other goes extinct. Finally, if both vaccine reproductive numbers are greater than one, coexistence of both diseases ensues. In Appendix B.3 we present numerical simulations in support of our results of this section.

**Figure 10:**
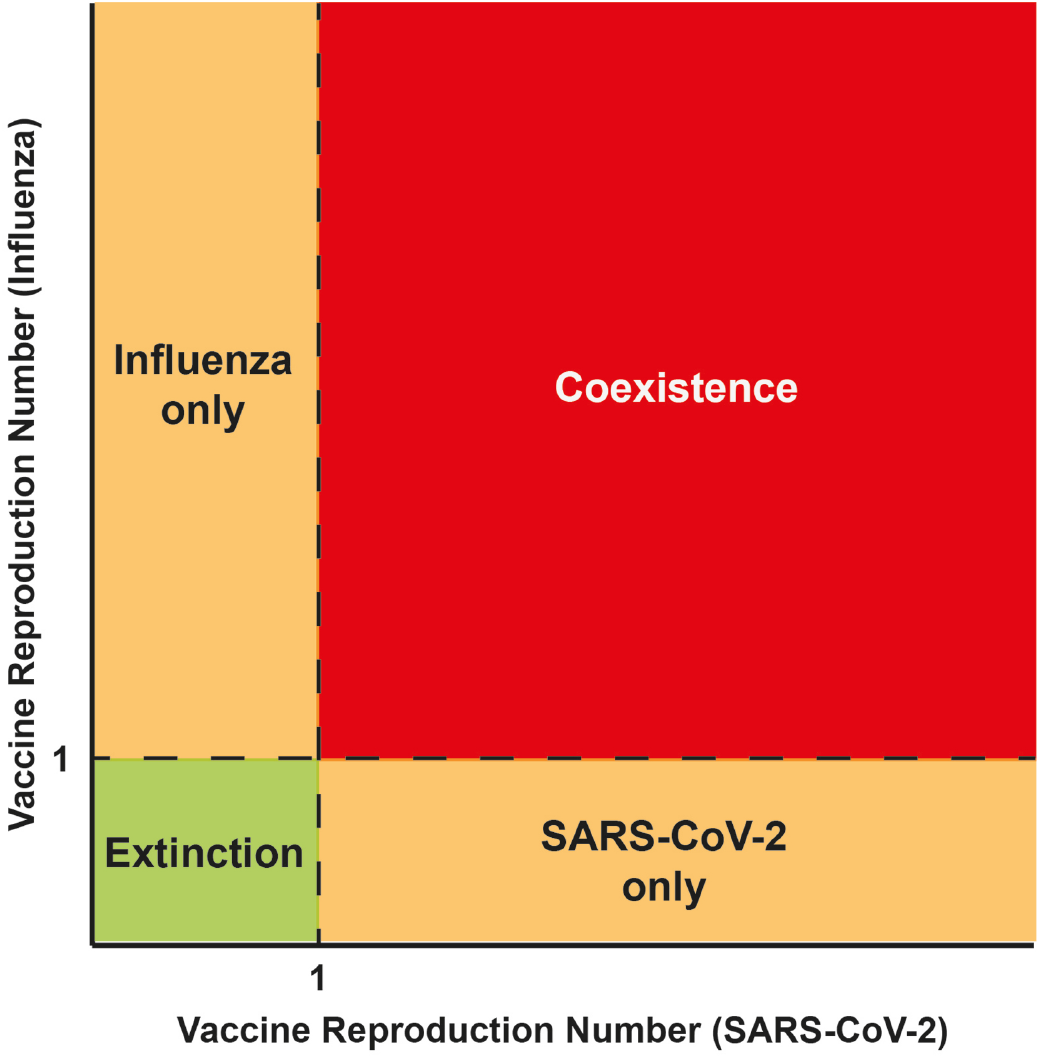
Summary of the asymptotic dynamics when varying both vaccine reproduction numbers. Scenarios for SARS-CoV-2 and influenza when *R*_*νk*_ *< R*_0*k*_, with *k* = *i, y*.

### 4.3 Seasonal contact rates

Finally, we address the issue of the long-term dynamics of the interactions between the two viruses. Seasonal variability is important to explain intra-annual fluctuations of viral populations [6, 7]. We incorporate seasonality using a periodic effective contact rate *β*_*k*_(*t*) = *β*_*k*_(1 + *ϵ* cos *ωt*) where 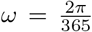 for an annual period; *β*_*k*_ is the baseline constant effective contact rate for virus *k* and *ϵ* is the amplitude of the seasonal variation (strength of the periodic forcing, 0 *< ϵ <* 1). Due to the higher reproduction number, the lack of previous immunity, the absence of antivirals or scarce vaccines, the equal and homogeneous effect of NPIs on reducing the effective contact rate, henceforth we consider SARS-CoV-2 as the competitively dominant virus in this interaction with influenza. This behavior is similar to the RSV over influenza.

Figure 11 illustrates alternation patterns between influenza and SARS-CoV-2. Here, parameter values are *β*_*i*_ = 0.45, *β*_*y*_ = 0.22, *α* = 0, *θ*_*i*_ = 1*/*365, *θ*_*y*_ = 1*/*180, *θ* = 1*/*365, *η*_*i*_ = 1*/*5, *η*_*y*_ = 1*/*14, *ω*_*i*_ = 1*/*5, *ω*_*y*_ = 1*/*14, *p*_*i*_ = 0.5, *E* = 0.8, *q*_*i*_ = 0.2, *q*_*y*_ = 0.3. Figure 11A shows that influenza and SARS-CoV-2 have stronger outbreaks every two years in an alternating sequence. This behavior is related to a reduction in susceptibility and an increase in the strength of the periodic forcing under a vaccination scheme. Note also that SARS-CoV-2 peaks are weaker every two years when they coincide with stronger influenza outbreaks and vice versa, suggesting competition for hosts.

**Figure 11:**
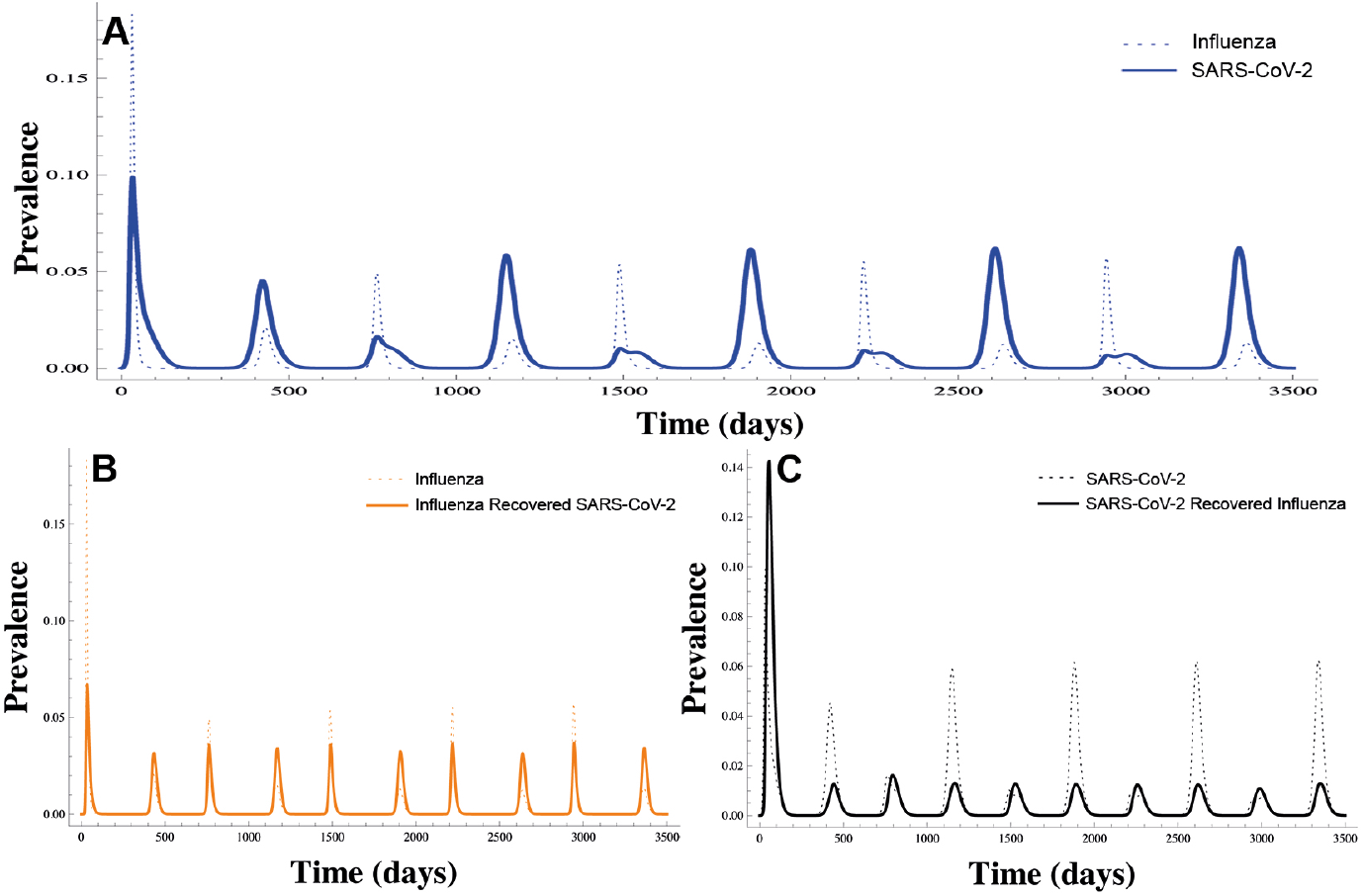
Alternation patterns. A) Annual prevalence cycles for primary influenza infections (blue line) and primary SARS-CoV-2 infections (dashed blue line). B) Annual cycles for primary (orange dashed line) and secondary (orange line) influenza infections. C) Annual cycles for primary (black dashed line) and secondary (black line) SARS-CoV-2 infections.

Figs. 11B-C show that this alternating sequence can be preserved even under vaccination for both viruses. Thus, in this scenario, every two years, the amplitude of the primary infections of both viruses is alternatively greater and, also, that this behavior may indeed correspond to the competitive alternation of both viruses.

Other patterns, between both viruses, are shown in Section B.4.

## 5 Conclusions

Influenza and SARS-CoV-2 will likely be co-circulating in the near future in many countries. However, vaccination campaigns have not begun at the same time. For example, in the Northern hemisphere the influenza vaccination campaign started in the Fall 2020, but that for SARS-CoV-2 has begun in early 2021 in many part of the World. Therefore, it is important to carefully plan vaccination campaigns and to define a realistic and sufficient coverage to avoid the situation described in the first paragraphs of this section.

Our results show that the parameters related to SARS-CoV-2, such as the average time of loss immunity, effective target coverage, and protective effect against infection by the coronavirus, all are relevant in reducing the vaccine reproduction number (Figure 9). To date, there are some parameter estimates for SARS-CoV-2 that remain unknown like temporary immunity and the large uncertainity of vaccine availability for developing or poor countries.

Our model also shows, as expected, that asymptotic behavior is closely associated with the vaccine reproduction number for each type of virus. For example, when considering realistic parameters for influenza and SARS-CoV-2, Figure 10 shows that if the vaccine reproduction number for influenza is greater than one and the vaccine reproduction number for SARS-CoV-2 is less than one, then influenza persists, and SARS-CoV-2 is eradicated. Coexistence sets in when both vaccine reproductive numbers are above one. This may happen as a consequence, for example, of low coverage or the time horizon for achieving it is large.

We have also numerically explored the behavior of our model with time-dependent effective contact rates. We consider it relevant because COVID-19 is a new disease with a transmission route similar to other viral infections, and in consequence, seasonal variability can explain future behaviors when considering co-circulation dynamics.

In general, we observe that influenza epidemics have less amplitude and show inter-epidemic periods with very low prevalence, whereas SARS-CoV-2 epidemics are broader in amplitude and show a clear endemic phase between outbreaks. In the simulated scenarios, we have observed that the prevalence of secondary cases (hosts that are susceptible to one but have recovered from the other virus) of both viruses decreases. The simulations assume an effective contact rate for influenza higher than that of SARS-CoV-2; however, the force of infection of this last virus (the one with the higher reproduction number) is much greater since the infection rate depends on the number of contacts per unit time, but also on the infection probability per contact and the infectious period which are different for both viruses. Also, we have found that the transmission patterns lead to alternation of patterns where influenza and SARS-CoV-2 have stronger outbreaks every two years in an alternating sequence, suggesting competition for hosts (see Figure 11).

In a more theoretical and general case, our results show that the vaccine reproduction number for some parameter combinations can be higher than the basic reproduction number opening up the possibility of the undesired outcome where vaccination may have a negative public health impact than otherwise at the population level. This outcome underlines the importance of the design of the vaccination strategy and of the availability of vaccines. Figure 2 shows that a low ETC may push the vaccine reproduction number above the basic reproduction number, resulting in a higher prevalence than the case without vaccination. This low ETC case is unrealistic in most contexts but could be of consequence in critical settings such as war, social disturbance, and natural disasters where coverage may fall short of the desired target.

Finally, in the absence of vaccination, we have shown that there are conditions under which the basic reproductive numbers do not need to be greater than one for both diseases to coexist (Figure 4). The above confirms the existence of bistability and of a backward bifurcation where for this special case, the recovery rate from the second infection for both viruses is slower than the recovery rate of the first infection. For this same situation, when *R*_0*k*_ *<* 1 *k* = *i, y*, there are initial conditions where superinfection can switch the stability of equilibria: low *α* gives coexistence and higher *α*, extinction (see Figure 7).

## Data Availability

In this work, we not consider data (therefore there is no statement regarding the availability)

## Acknowledgments

All authors acknowledge support from DGAPA-PAPIIT-UNAM grant IV100220 (convocatoria especial COVID-19), JXVH acknowledges support from DGAPA-PAPIIT-UNAM grant IN115720. MAAZ acknowledges support from PRODEP Programme (No. 511-6/2019-8291). MNL acknowledges support from the Asociación Mexicana de Cultura, A.C.

## Competing Interests

The authors declare no conflicts of interest.

## Authors Contributions

MAAZ, MNL, MSC and JXVH conceived and formulated the problem, MAAZ, MNL and JXVH performed analysis and simulations. MSC performed the sensitivity analysis. MAAZ, MNL, MSC and JXVH wrote the paper. All authors discussed the results and conclusions.

## Appendix A Local stability of disease-free equilibrium

To prove lemma 3.3, we compute the jacobian matrix of eq. (1) and evaluate it at disease-free equilibrium *E*_0_ (eq. (3)). Thus:

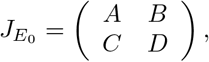

where,

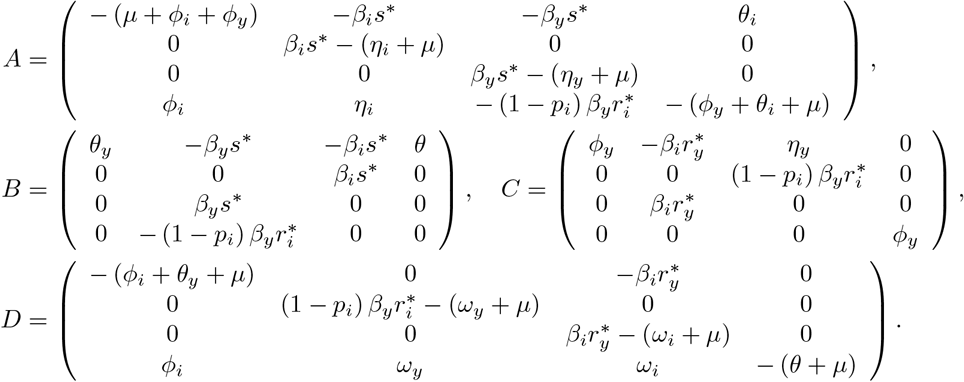

Then, the characteristic polinomial of the 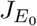 is

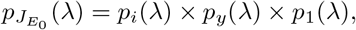

with

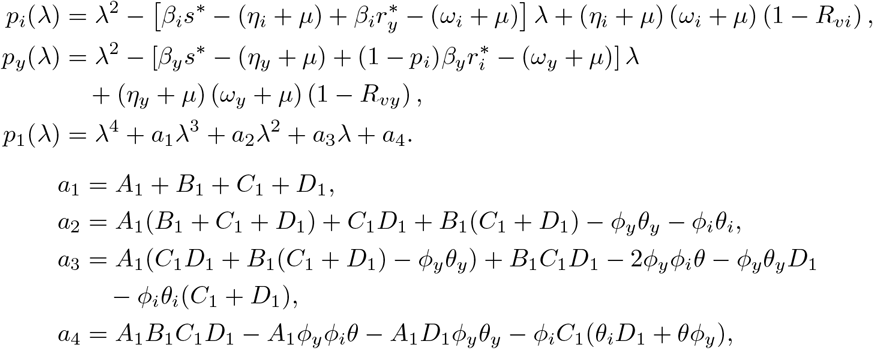

where

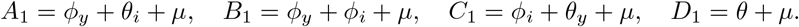

We observe that *a*_1_, *a*_2_, *a*_3_, *a*_4_ *>* 0 and 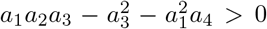. Hence, *p*_1_(*λ*) satisfying the RouthHurwitz criterion. In consequence, all roots of *p*_1_(*λ*) have negative real parts. Likewise, it is clear that all roots of *p*_*k*_(*λ*) are negative if and only if *R*_*νk*_ *<* 1, with *k* = *i, y*. Therefore, all eigenvalues of 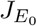 have negative real parts if and only if *R*_*ν*_ *<* 1.

## Appendix B Influenza and SARS-CoV-2

### B.1 Model parametrization

To explore numerical scenarios from Influenza and SARS-CoV-2, we estimate the baseline parameters from bibliographical sources. According to [32], for seasonal influenza the median reproduction number is 1.28 and that for SARS-CoV-2 is 2.79, but sources report large variability [33]. Likewise, [10] citing several sources, reports an *R*_0_ for influenza in the range 1.06–3.4 with a mean of 1.68. The same source gives an incubation period in the range of 1–6.3 days with a mean of 2.61 and an infectious period (*η*_*i*_) range of 1–9 days with a mean of 4.58 days. For SARS-CoV-2, [34] reports an incubation period of 3 to 4 days and an infectious period (*η*_*y*_) of 4–5 days but [10] gives an incubation period in the range 1.9 to 14.7 days with a mean of 5 days, and an infectious period in the range 7–35 days with a mean of 15.2 days.

Influenza vaccine efficacy varies every year. For 2019–2020 the [35] reports *σ*_*i*_ = 0.29 but in 2010-2011 *σ*_*i*_ = 0.6. [36, 37] have argued that efficacy declines because of waning immunity that may last 6 months for influenza A(H1N1) and influenza B and at least 5 months for influenza A(H3N2). Besides this reported efficacies we are postulation a parameter *p*_*i*_ that mimics a protective role conveyed by influenza vaccination against SARS-CoV-2 infection. This hypothesis is based on the work of [38] that reports that protective influenza vaccination does not negatively affect the risk of contracting coronaviruses. We explore the possibility that the effect is positive, thus conferring a reduction in the risk of SARS-CoV-2 infection.

For SARS-CoV-2, several vaccines have been deployed with efficacies in the range of 50-95% with more likely scenarios of 70%. The Pfizer, Moderna vaccines have efficacies at the upper end of this interval. Astra-Zeneca vaccine efficacy is around 75%. On the other hand, coverage has three basic scenarios: low 20%, medium 50%, and high 80%. Given the form in which we are modeling coverage, we set up scenarios where the above percentages are reached after *T* = 90, 180 and 365 days. We assume that SARS-CoV-2 immunity ranges from half a year to lifelong, with a more likely scenario of one year. These estimates are largely based on data on immunity to other coronaviruses [39]. Currently, whether past infections will prevent severe COVID-19 on re-infection to SARS-CoV-2 is not known.

### B.2 Reproduction numbers, vaccine efficacy, effective coverage and temporary immunity

Figure B.2.1: behavior of the vaccine reproduction number (purple surface) and vaccine reproduction number for SARS-CoV-2 (orange surface) for, *θ*_*y*_ = 1*/*180, vaccine efficacy is 95% (Figure B.2.1A) and target coverage is 20% (Figure B.2.1B). Other parameters as in Figure 9. Both figures show the importance of a quick vaccination campaign (time horizon close to zero) to effectivelly reduce *R*_*νy*_. Figure B.2.1A: there is a threshold value of target coverage after which *R*_*ν*_ is constant. (*R*_*νi*_ does not depend on target coverage and time horizon for SARS-CoV-2 vaccination). Figure B.2.1B shows a similar behavior for SARS-CoV-2 vaccination decreases.

### B.3 Transmission reduction by vaccination (*R*_*νk*_ *< R*_0*k*_)

To illustrate some specific scenarios of Section 4.2, we fix *µ* = 0.000039139, *α* = 0, *η*_*i*_ = 1*/*5, *η*_*y*_ = 1*/*14, *θ* = 1*/*365, *θ*_*i*_ = 1*/*365, *ω*_*i*_ = 1*/*5, *ω*_*y*_ = 1*/*14, *p*_*i*_ = 0.5, *ϕ* _*i*_ = 0.001603, *ϕ* _*y*_ = 0.002341 and *θ*_*y*_ = 1*/*180. The effective contact rates are constant but different in each scenario.

Figure B.3.1 shows SARS-CoV-2 persistence. *β*_*i*_ and *β*_*y*_ are 0.3 and 0.2, respectively, giving *R*_*νi*_ *<* 1 *< R*_*νy*_. In Figure B.3.1A red line represents dynamics under vaccination. Blue lines, without vaccination. Vaccine coverage reduces the prevalence of SARS-CoV-2, with the extinction of influenza (Figure B.3.1A). Influenza persists and SARS-CoV-2 goes extinct when *R*_*νy*_ *<* 1 *< R*_*νi*_.

**Figure B.2.1:**
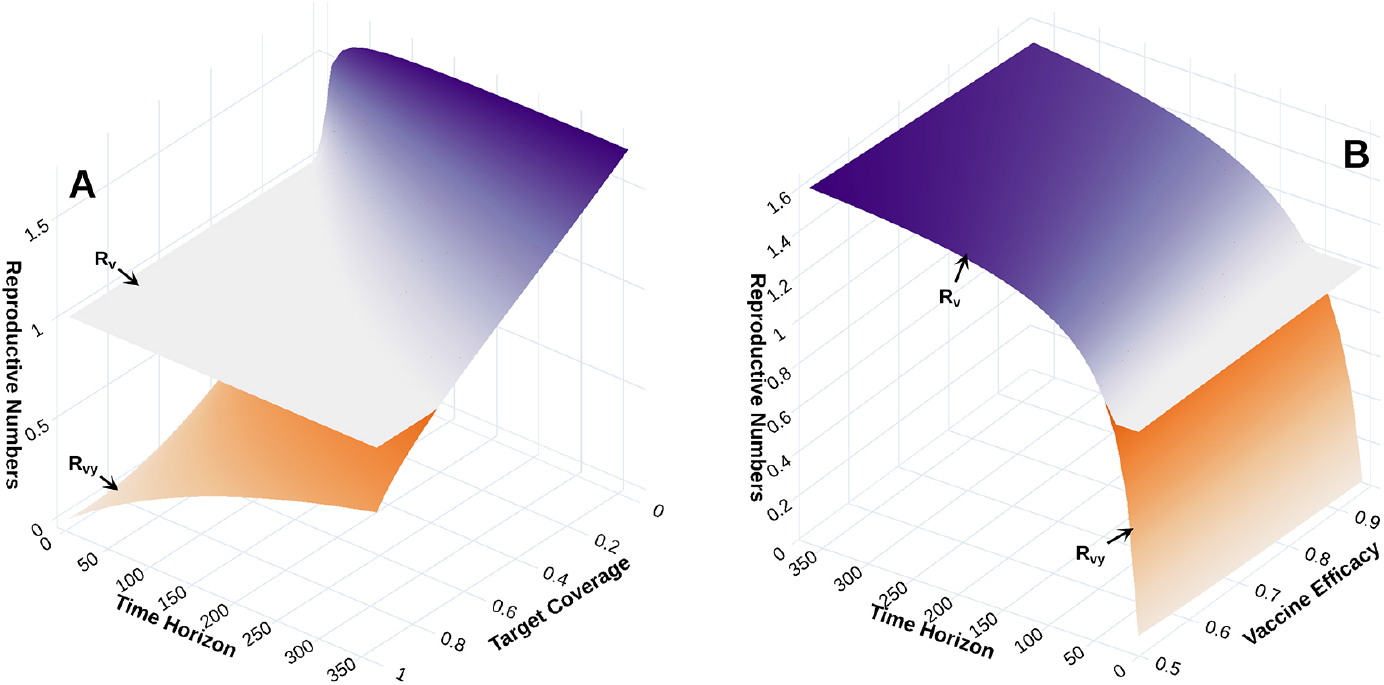
Reproduction number behavior. A) Reproduction numbers as a function of target coverage and time horizon for SARS-CoV-2 vaccination. B) Reproduction numbers as a function of vaccine efficacy and time horizon for SARS-CoV-2 vaccination. Target coverage for influenza is 35% in 4 months with a vaccine efficacy equal to 50%.

**Figure B.3.1:**
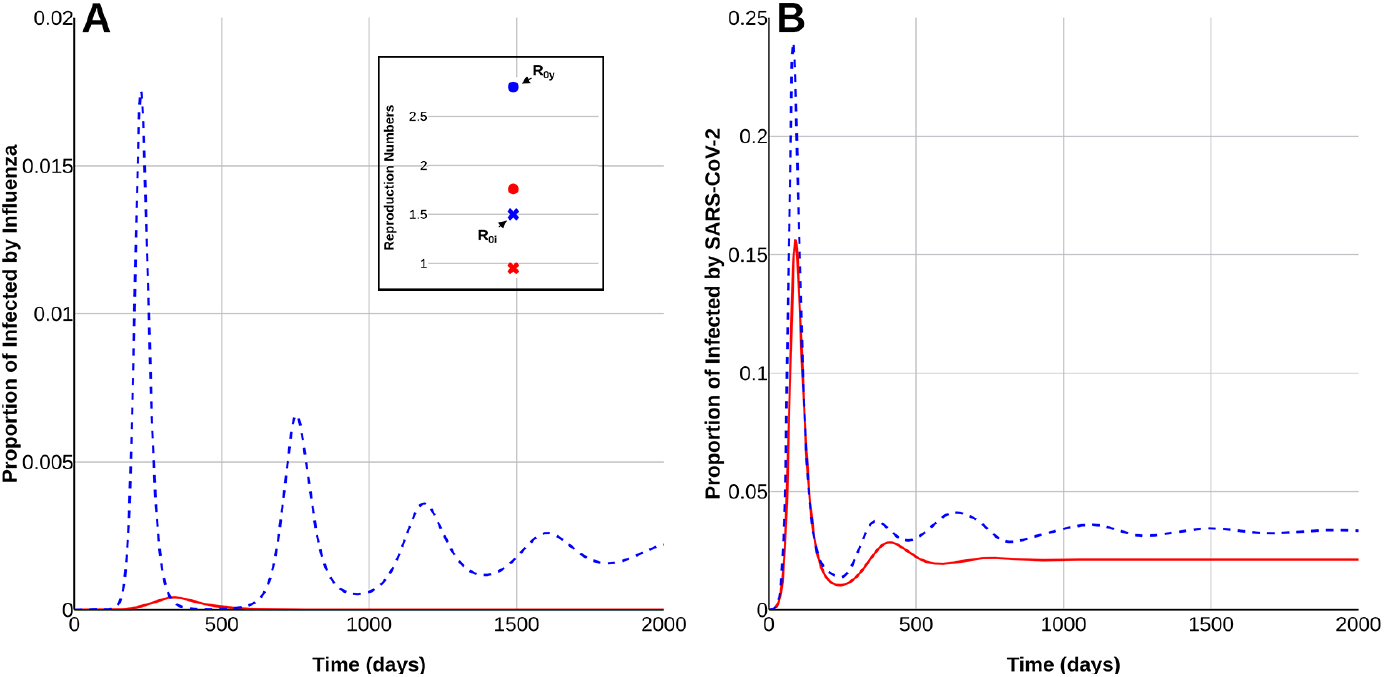
Extinction of influenza and persistence of SARS-CoV-2 with simultaneous vaccination. In the inset dots represent SARS-CoV-2 reproduction numbers, and crosses influenza reproduction numbers. Color codes: Red vaccination is applied, blue, no vaccination. A) Proportion of individuals infected with influenza. B) Proportion of individuals infected with SARS-CoV-2. Solid red lines represent disease dynamics when vaccination for both viruses is simultaneous. Dashed blue lines show dynamics without vaccination.

Figure B.3.2: *β*_*i*_ and *β*_*y*_ are 0.35 and 0.2, so both reproductive numbers greater than one. Endemic equilibrium levels in the presence of vaccination are reduced compared to the no vaccination case (dashed blue lines).

**Figure B.3.2:**
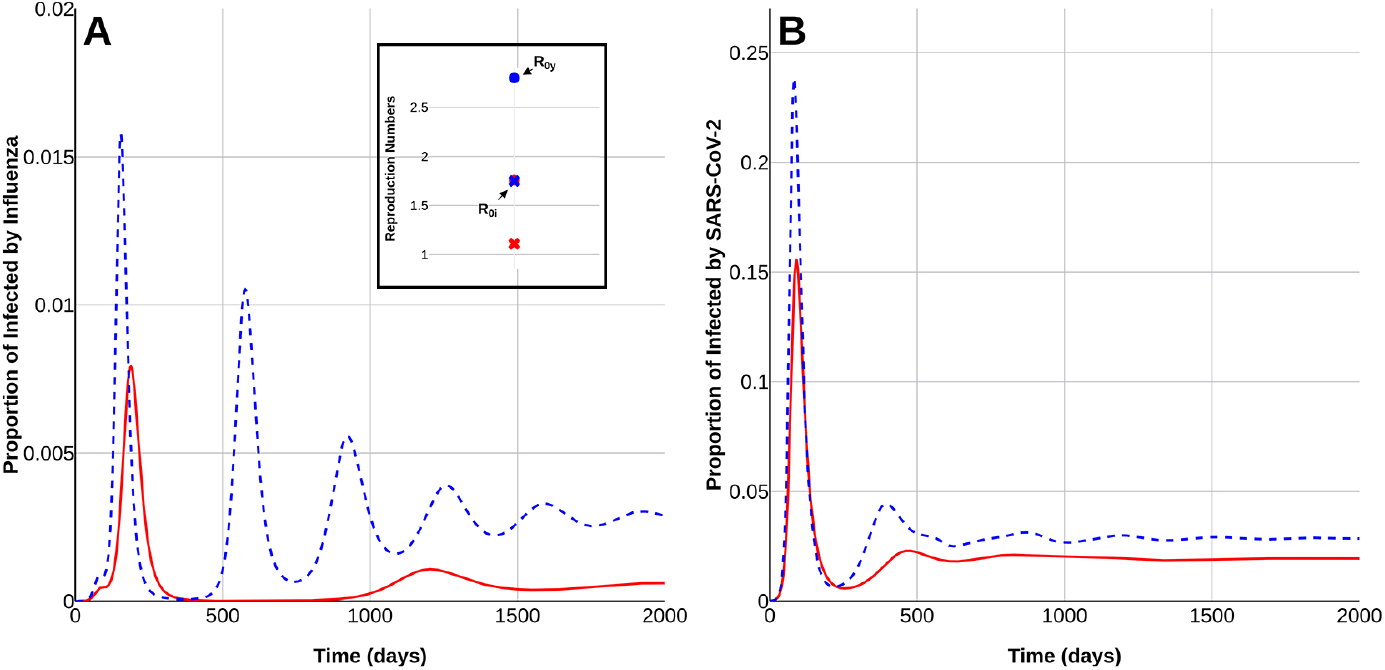
Coexistence of influenza and SARS-CoV-2. A) Proportion of infected individuals with influenza. B) Proportion of infected individuals with SARS-CoV-2. Solid red lines: vaccine application. Dashed blue lines: no vaccination.

### B.4 Seasonal contact rates

Figure B.4.1: alternating outbreaks. *β*_*i*_ = 0.4, *β*_*y*_ = 0.2, *α* = 0, *θ*_*i*_ = 1*/*365, *θ*_*y*_ = 1*/*365, *θ* = 1*/*365, *η*_*i*_ = 1*/*5, *η*_*y*_ = 1*/*14, *ω*_*i*_ = 1*/*5, *ω*_*y*_ = 1*/*14, *p*_*i*_ = 0.5, *E* = 0.9. Figure B.4.1A: time trajectory of both viruses with vaccination. Outbreaks alternate in time with influenza oscillations presenting narrower amplitudes. In contrast, SARS-CoV-2 outbreaks are broader. Both present inter-epidemic periods with very low prevalence. Epidemic outbreaks do not overlap. In secondary infections the alternation of patterns persists but prevalence of both viruses decreases (see Figs. B.4.1B-C): the effect of the vaccine is observed in SARS-CoV-2 and influenza, according to the magnitude of the first outbreak of primary and secondary infections of both viruses.

Every year influenza arrives in waves with each new influenza epidemic produced, in general, by a different viral strain in a process of lineage or variant replacement [40]. To mimic this situation in a simple, yet reasonable way, we aggregate all the influenza strains as a single influenza epidemic and allow for reinfections given the assumption that temporary immunity against influenza lasts one year. With this simplification, our model produces an annual pattern driven by the yearly weather variability.

Figure B.4.2A: Vaccination induces a synchronized oscillatory pattern in both viruses now strongly associated to the strength of the periodic forcing. This periodic behavior is also reflected in secondary infections (see Figs. B.4.2B-C). For this scenario, parameter values are *β*_*i*_ = 0.48, *β*_*y*_ = 0.25, *α* = 0, *θ*_*i*_ = 1*/*365, *θ*_*y*_ = 1*/*180, *θ* = 1*/*365, *η*_*i*_ = 1*/*5, *η*_*y*_ = 1*/*14, *ω*_*i*_ = 1*/*5, *ω*_*y*_ = 1*/*14, *p*_*i*_ = 0.5, *ϵ* = 0.5 *q*_*i*_ = 0.2, *q*_*y*_ = 0.3.

### B.5 Sensitivity analysis

For completeness, a variance based sensitivity analysis known as Sobol method [41, 42] was conducted to evaluate the parameters’ influence on the model’s state variables. A global sensitivity analysis assumes that the output of a system is a function of a set of inputs (parameters). By assuming that the vector of parameters is a random variable, the output is also a random variable. The total variability of the output, induced by the variability of the inputs, is decomposed in proportions associated with individual or sets of parameters. The higher the proportion of variability caused by changes in a specific parameter, then the higher the sensibility of the model to that parameter. *θ, θ*_*i*_, *θ*_*y*_, *ϕ*_*i*_, *ϕ*_*y*_, *p*_*i*_, *β*_*i*_, *β*_*y*_ vary uniformly in the ranges presented in Table B.5.1, while parameters *η*_*i*_ = 1*/*5, *η*_*y*_ = 1*/*14, *ω*_*i*_ = 1*/*5 and *ω*_*y*_ = 1*/*14 are held constant.

**Figure B.4.1:**
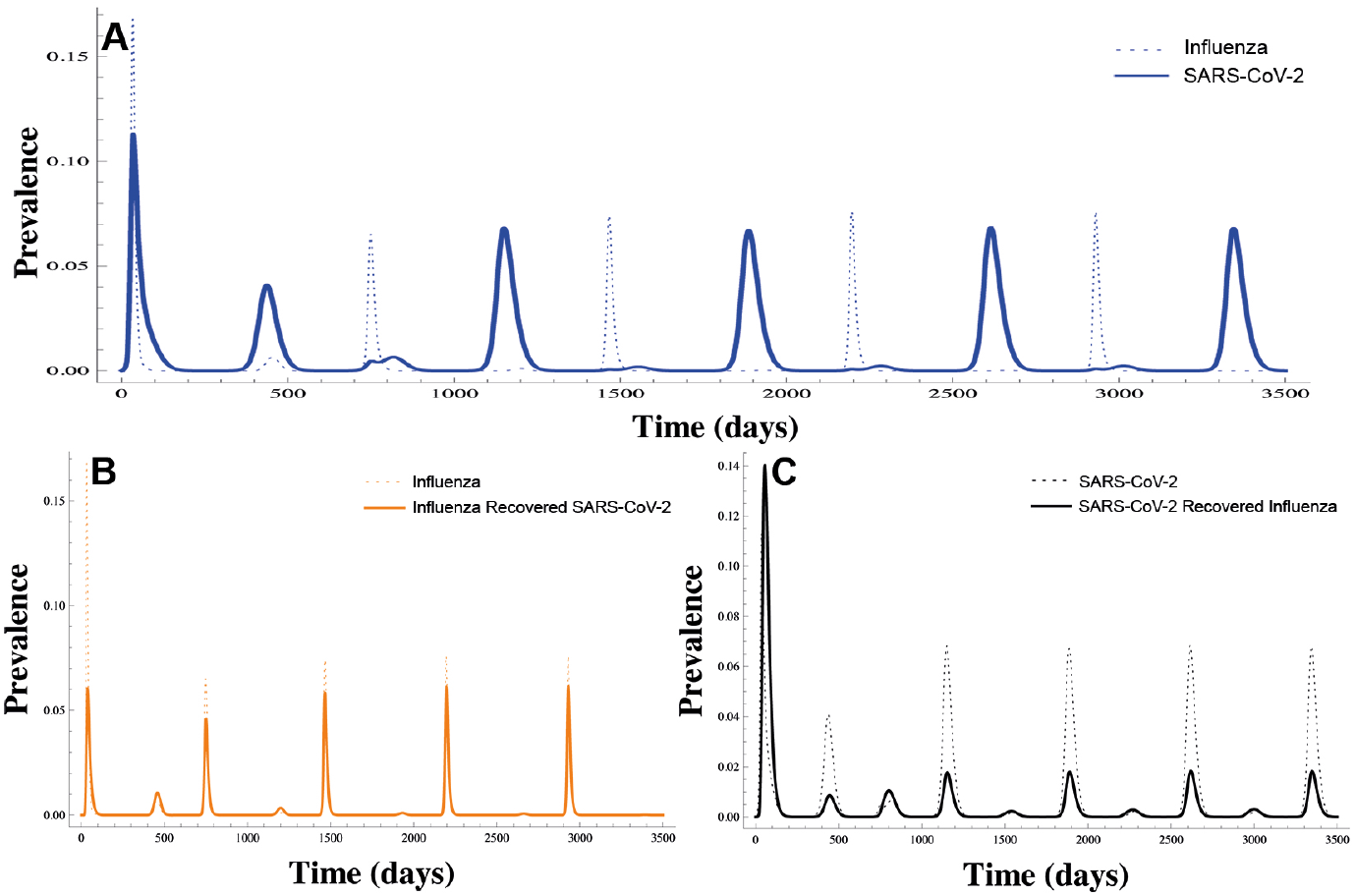
Alternating outbreaks. A) Numerical simulation of primary influenza infections (blue dashed line) and primary SARS-CoV-2 infections (blue line). B) Primary (dashed orange line) and secondary influenza (orange line) infections. C) Primary (dashed black line) and secondary SARS-CoV-2 infections (black line).

**Table B.5.1:** Parameter ranges for the sensitivity analysis.

| Parameter | Lower bound | Upper bound |
| --- | --- | --- |
| $p_i$ | 0 | 1 |
| $\beta_i$ | 0 | 0.4 |
| $\beta_y$ | 0 | 0.3 |
| $\phi_i$ | 0 | 1/180 |
| $\phi_y$ | 0 | 1/365 |
| $\theta$ | 1/540 | 1/180 |
| $\theta_i$ | 1/540 | 1/180 |
| $\theta_y$ | 1/365 | 1/180 |

Figure B.5.1: Sobol indices for primary infections of both virus. For influenza (*i*), *β*_*i*_ is the most important parameter, with its individual influence decreasing over time. *ϕ*_*i*_ is the second most influential parameter followed by *β*_*y*_. This implies that changes in vaccination schemes are indeed important to control influenza. In the case of the SARS-CoV-2 (*y*) virus, the most dominant parameter is *β*_*y*_ while the influence of the others is very small. Figure B.5.2: Sobol indices for secondary infections. It can be seen that the influence of the parameters on primary and secondary infections of influenza are very similar.In the case of SARS-CoV-2 secondary infections, the protective effect gained from influenza (*p*_*i*_) is only below *β*_*y*_ in terms of importance.

**Figure B.4.2:**
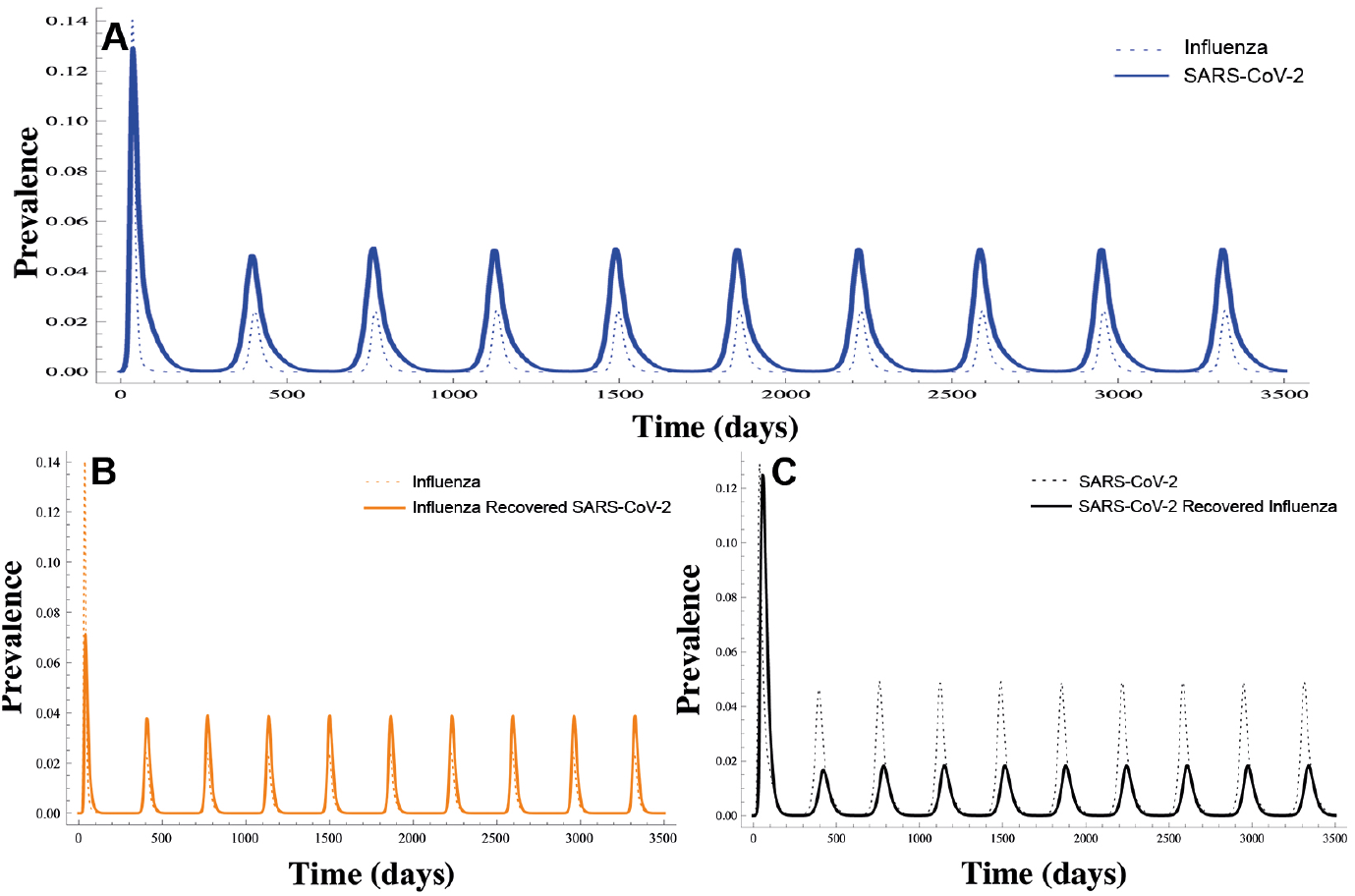
Synchronized outbreak patterns. A) Annual prevalence cycles for primary influenza infections (blue dashed line) and primary SARS-CoV-2 infections (blue line) B) Annual cycles for primary (orange dashed line) and secondary (orange line) influenza infections. C) Annual cycles for primary (black dashed line) and secondary (black line) SARS-CoV-2 infections.

We also perform a sensitivity analysis for the vaccine reproduction numbers *R*_*νi*_ and *R*_*νy*_, Figure B.5.3. For each disease, the parameter that changes the most the reproduction number is the corresponding contact rate. The other parameters that show an important effect are the vaccination coverage *ϕ*_*i*_ and *ϕ*_*y*_ which, of course, depend on the corresponding ETC 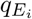 and 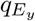.

**Figure B.5.1:**
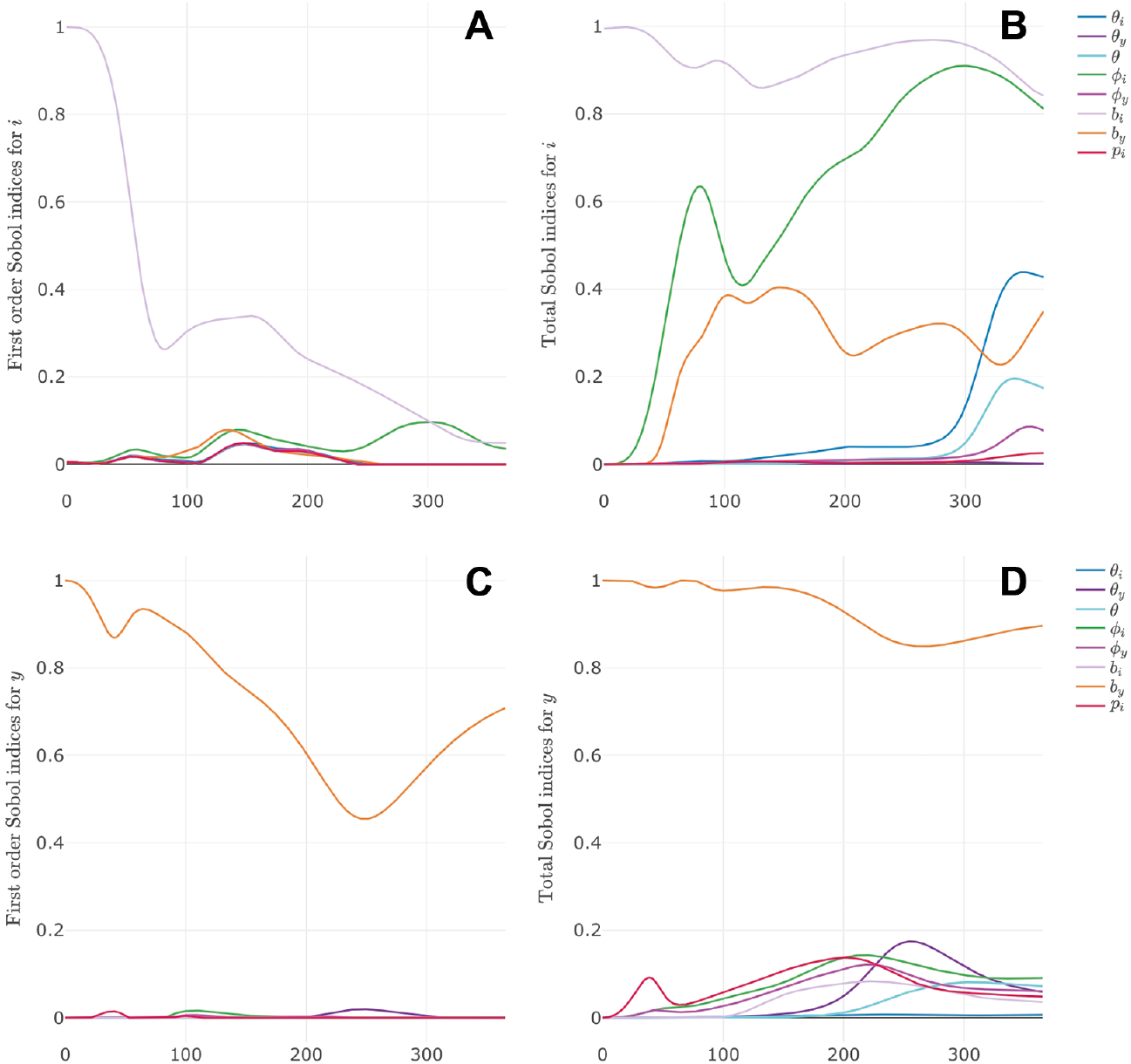
Sensitivity analysis. Left panels show the first order Sobol indices for the individual effects on influenza prevalence *i* (upper left panel) and SARS-CoV-2 prevalence *y* (lower left panel) at each point in time. Right panels show total Sobol indices, for the effect of each parameter and its interactions with all other parameters.

**Figure B.5.2:**
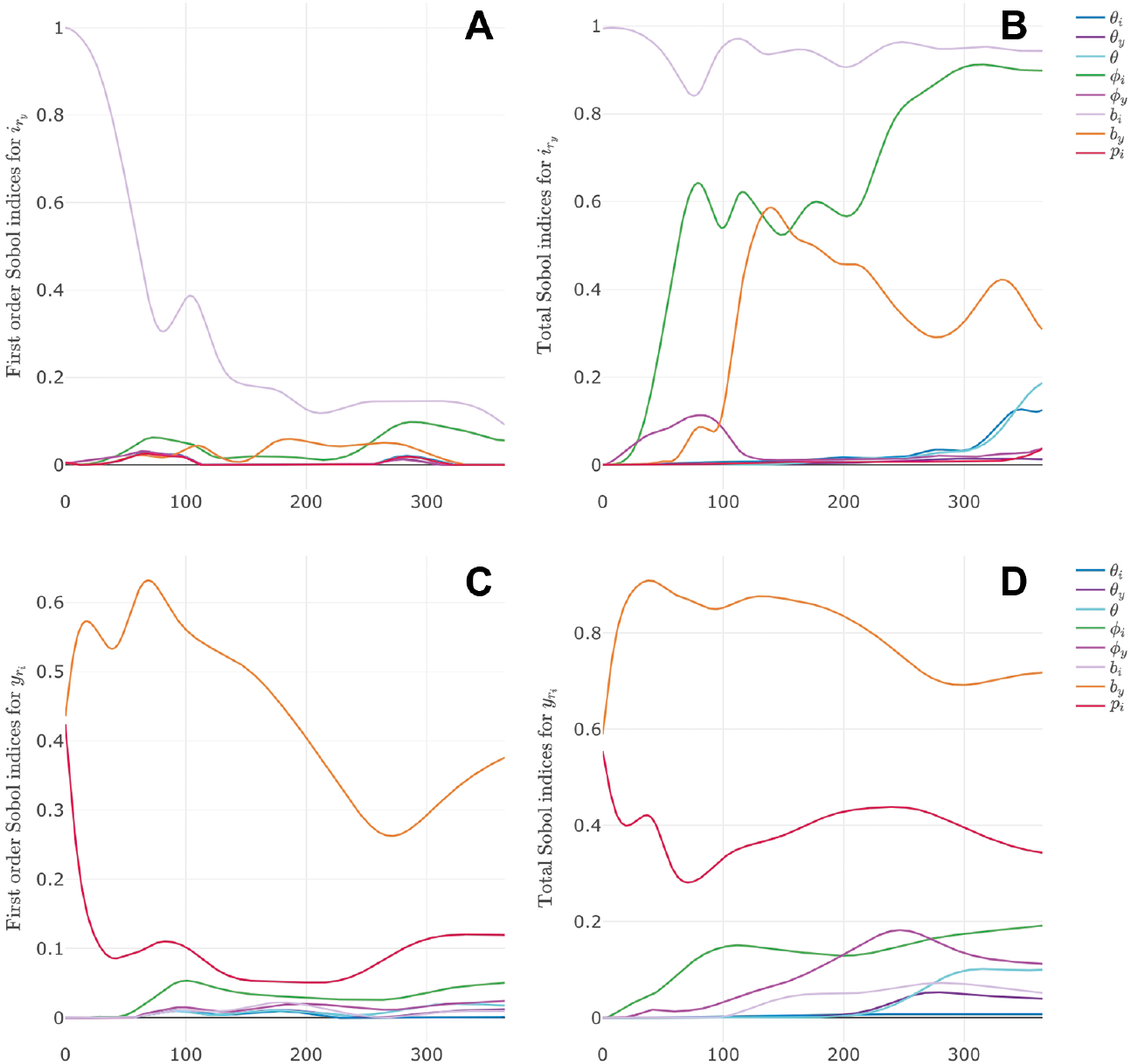
Sensitivity analysis. Left panels show the first order Sobol indices for the individual effects on secondary influenza prevalence 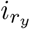 (upper left panel) and secondary SARS-CoV-2 prevalence 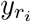 (lower left panel) at each point in time. Right panels show total Sobol indices, for the effect of each parameter and its interactions with all other parameters.

**Figure B.5.3:**
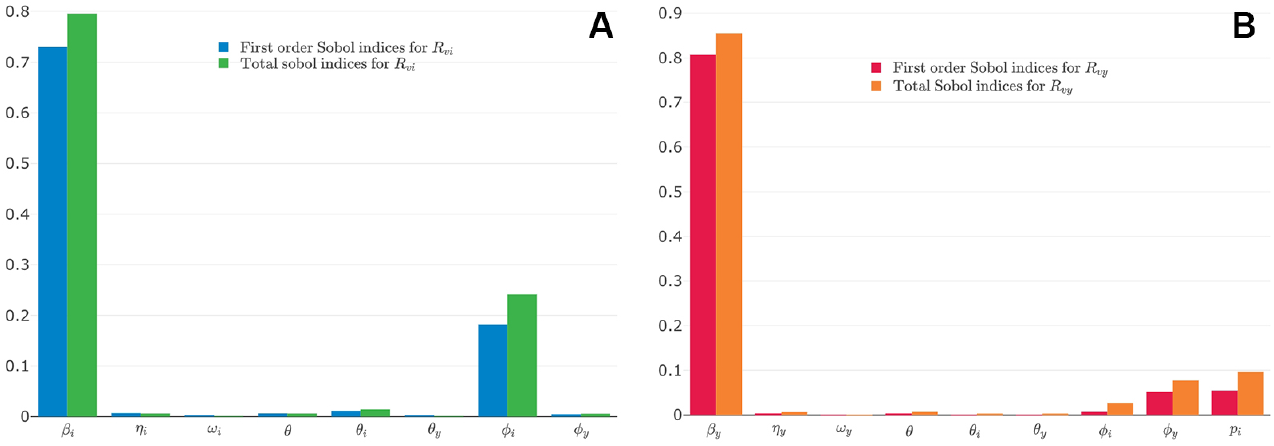
Sensitivity analysis for the vaccine reproduction numbers for influenza *R*_*νi*_ (A) and SARS-CoV-2 *R*_*νy*_ (B).

## Notes

### Competing Interest Statement

The authors have declared no competing interest.

### Clinical Trial

Clinical trials were not required for this work

### Author Declarations

For this work we do not consider IRB

### Summary of Updates

Mathematical model set-up updated to clarify Section 3; Supplemental files updated

## References

[1] S. Levin and D. Pimentel, “Selection of Intermediate Rates of Increase in Parasite-Host Systems,” The American Naturalist, vol. 117, no. 3, pp. 308–315, 1981.

[2] H. Bremermann and H. Thieme, “A competitive exclusion principle for pathogen virulence,” Journal of Mathematical Biology, vol. 27, pp. 179–190, Apr. 1989.

[3] M. A. Nowak and R. M. May, “Superinfection and the Evolution of Parasite Virulence,” Proceedings of the Royal Society B: Biological Sciences, vol. 255, no. 1342, pp. 81–89, 1994.

[4] C. Castillo-Chavez and J. Velasco-Hernández, “On the Relationship Between Evolution of Virulence and Host Demography,” Journal of theoretical Biology, vol. 192, pp. 437–444, jun 1998.

[5] D. Tilman, “Competition and Biodiversity in Spatially Structured Habitats,” Ecology, vol. 75, no. 1, pp. 2–16, 1994.

[6] J. X. Velasco-Hernández, M. Núñez-López, A. Comas-García, D. E. N. Cherpitel, and M. C. Ocampo, “Superinfection between Influenza and RSV Alternating Patterns in San Luis Potos State, Mxico,” PLOS ONE, vol. 10, pp. 1–19, 03 2015.

[7] C. Johnston, A. Magaret, P. Roychoudhury, A. L. Greninger, D. Reeves, J. Schiffer, K. R. Jerome, C. Sather, K. Diem, J. R. Lingappa, C. Celum, D. M. Koelle, and A. Wald, “Dual-strain genital herpes simplex virus type 2 (HSV-2) infection in the US, Peru, and 8 countries in sub-Saharan Africa: A nested cross-sectional viral genotyping study,” PLoS Medicine, vol. 14, no. 12, pp. 1–17, 2017.

[8] R. M. May and M. A. Nowak, “Superinfection, metapopulation dynamics, and the evolution of diversity,” Journal of Theoretical Biology, vol. 170, no. 1, pp. 95–114, 1994.

[9] R. Levins and D. Culver, “Regional Coexistence of Species and Competition between Rare Species,” Proceedings of the National Academy of Sciences, vol. 68, no. 6, pp. 1246–1248, 1971.

[10] J. Spencer, D. P. Shutt, S. K. Moser, H. Clegg, H. J. Wearing, H. Mukundan, and C. A. Manore, “Epidemiological parameter review and comparative dynamics of influenza, respiratory syncytial virus, rhinovirus, human coronavirus, and adenovirus,” medRxiv, 2020.

[11] L. G. Gallo, A. Flávia, D. M. Oliveira, and A. A. Abrahão, “Ten Epidemiological Parameters of COVID-19 : Use of Rapid Literature Review to Inform Predictive Models During the Pandemic,” Frontiers in Public Health, vol. 8, p. 830, 2020.

[12] J. Sultana, G. Mazzaglia, N. Luxi, A. Cancellieri, A. Capuano, C. Ferrajolo, C. de Waure, G. Ferlazzo, and G. Trifir, “Potential effects of vaccinations on the prevention of COVID-19: rationale, clinical evidence, risks, and public health considerations,” Expert Review of Vaccines, vol. 19, no. 10, pp. 919–936, 2020.

[13] K. M. Bubar, S. M. Kissler, M. Lipsitch, S. Cobey, Y. H. Grad, and D. B. Larremore, “Model-informed COVID-19 vaccine prioritization strategies by age and serostatus,” medRxiv, 2020.

[14] Z. McCarthy, S. Athar, M. Alavinejad, C. Chow, I. Moyles, K. Nah, J. D. Kong, N. Agrawal, A. Jaber, L. Keane, S. Liu, M. Nahirniak, D. S. Jean, R. Romanescu, J. Stockdale, B. T. Seet, L. Coudeville, E. Thommes, A. F. Taurel, J. Lee, T. Shin, J. Arino, J. Heffernan, A. Chit, and J. Wu, “Quantifying the annual incidence and underestimation of seasonal influenza: A modelling approach,” Theoretical Biology and Medical Modelling, vol. 17, no. 1, pp. 1–16, 2020.

[15] R. E. Baker, A. S. Mahmud, C. E. Wagner, W. Yang, V. E. Pitzer, C. Viboud, G. A. Vecchi, C. J. E. Metcalf, and B. T. Grenfell, “Epidemic dynamics of respiratory syncytial virus in current and future climates,” Nature Communications, vol. 10, no. 1, pp. 1–8, 2019.

[16] V. E. Pitzer, C. Viboud, W. J. Alonso, T. Wilcox, C. J. Metcalf, C. a. Steiner, A. K. Haynes, and B. T. Grenfell, “Environmental Drivers of the Spatiotemporal Dynamics of Respiratory Syncytial Virus in the United States,” PLOS Pathogens, vol. 11, pp. 1–14, jan 2015.

[17] Conlon, C. Ashur, L. Washer, K. K.A., and M. Hofmann Bowman, “Impact of the influenza vaccine on COVID-19 infection rates and severity,” American Journal of Infection Control, 2021.

[18] Patwardhan and A. Ohler, “The Flu Vaccination May Have a Protective Effect on the Course of COVID-19 in the Pediatric Population: When Does Severe Acute Respiratory Syndrome Coronavirus 2 (SARS-CoV-2) Meet Influenza?,” Cureus, vol. 13, p. e12533, 2021.

[19] H. L. Smith and H. R. Thieme, Dynamical systems and population persistence. American Mathematical Society, 2011.

[20] P. van den Driessche and J. Watmough, “Reproduction numbers and sub-threshold endemic equilibria for compartmental models of disease transmission,” Mathematical biosciences, vol. 180, pp. 29–48, 2002.

[21] M. Kribs-Zaleta and J. Velasco-Hernández, “A simple vaccination model with multiple endemic states,” Mathematical Biosciences, vol. 164, pp. 183–201, apr 2000.

[22] E. L. Allgower and K. Georg, Introduction to Numerical Continuation Methods. SIAM Classics in Applied Mathematics 45, 2003.

[23] F. Brauer, “Backward bifurcations in simple vaccination models,” J. Math. Anal. Appl., vol. 298, no. 2, pp. 418–431, 2009.

[24] O. Sharomi, C. Podder, G. A.B., E. Elbasha, and W. J., “Role of incidence function in vaccine-induced backward bifurcation in some HIV models,” Math. Biosci., vol. 210, no. 2, pp. 436–463, 2007.

[25] M. Lipsitch and M. M.B. Murray, “Multiple equilibria: tuberculosis transmission require un-realistic assumptions,” Theor. Popul. Biol., vol. 63, pp. 169–170, 2003.

[26] E. Elbasha, C. Podder, and G. A.B., “Analyzing the dynamics of an SIRS vaccination model with waning natural and vaccine-induced immunity,” Nonlinear Anal. RWA, vol. 12, no. 5, pp. 2692–2705, 2011.

[27] O. Sharomi and A. Gumel, “Reinfection-induced backward bifurcation in the transmission dynamics of Chlamydia trachomatis,” J. Math. Anal. Appl., vol. 356, pp. 96–118, 2009.

[28] J. Dushoff, H. Wenzhang, and C.-C. C., “Backwards bifurcations and catastrophe in simple models of fatal diseases,” J. Math. Biol., vol. 36, pp. 227–248, 1998.

[29] World Health Organization, “Influenza Update 380,” Tech. Rep. November, World Health Organization, 2020.

[30] J. P. A. Ioannidis, “Global perspective of COVID-19 epidemiology for a full-cycle pandemic,” European Journal of Clinical Investigation, vol. 50, no. 12, p. e13423, 2020.

[31] J. Jung, “Epidemiologic Evaluation and Risk Communication Regarding the Recent Reports of Sudden Death after Influenza Vaccination in the COVID-19 Pandemic,” Journal of Korean medical science, vol. 35, no. 41, p. e378, 2020.

[32] T. Chotpitayasunondh, T. K. Fischer, J. M. Heraud, A. C. Hurt, A. S. Monto, A. Osterhaus, Y. Shu, and J. S. Tam, “Influenza and COVID-19: What does co-existence mean?,” Influenza and other Respiratory Viruses, 2020.

[33] M. Biggerstaff, S. Cauchemez, C. Reed, M. Gambhir, and L. Finelli, “Estimates of the reproduction number for seasonal, pandemic, and zoonotic influenza: A systematic review of the literature,” BMC Infectious Diseases, vol. 14, no. 1, pp. 1–20, 2014.

[34] Y. M. Bar-On, R. Sender, A. I. Flamholz, R. Phillips, and R. Milo, “A quantitative compendium of COVID-19 epidemiology,” arXiv, 2020.

[35] Centers for Disease Control and Prevention, “CDC Seasonal Flu Vaccine Effectiveness Studies.” Accesed on December 3, 2020.

[36] J. M. Ferdinands, A. M. Fry, S. Reynolds, J. G. Petrie, B. Flannery, M. L. Jackson, and E. A. Belongia, “Intraseason waning of influenza vaccine protection: Evidence from the US influenza vaccine effectiveness network, 2011-2012 through 2014-2015,” Clinical Infectious Diseases, vol. 64, no. 5, pp. 544–550, 2017.

[37] J. M. Ferdinands, M. M. Patel, I. M. Foppa, and Alicia M. Fry, “Influenza vaccine effectiveness,” Clinical Infectious Diseases, vol. 69, no. 1, p. 190, 2019.

[38] M. Skowronski, M. Zou, Q. Clarke, C. Chambers, J. A. Dickinson, S. Sabaiduc, R. Olsha, J. B. Gubbay, S. J. Drews, H. Charest, A.-L. Winter, A. Jassem, M. Murti, M. Krajden, and G. De Serres, “Influenza Vaccine Does Not Increase the Risk of Coronavirus or Other Noninfluenza Respiratory Viruses: Retrospective Analysis From Canada, 2010?2011 to 2016?2017,” Clinical Infectious Diseases, vol. 71, no. 16, pp. 2285–2288, 2020.

[39] P. K. Kiyuka, C. N. Agoti, P. K. Munywoki, R. Njeru, A. Bett, J. R. Otieno, G. P. Otieno, E. Kamau, T. G. Clark, L. van der Hoek, P. Kellam, D. J. Nokes, and M. Cotten, “Human Coronavirus NL63 Molecular Epidemiology and Evolutionary Patterns in Rural Coastal Kenya,” The Journal of Infectious Diseases, vol. 217, pp. 1728–1739, 03 2018.

[40] K. Koelle, S. Cobey, B. Grenfell, and M. Pascual, “Epochal Evolution Shapes the Phylodynamics of Interpandemic Influenza A (H3N2) in Humans,” Science, vol. 314, no. 5807, pp. 1898–1903, 2006.

[41] Sobol, “Global sensitivity indices for nonlinear mathematical models and their Monte Carlo estimates,” Mathematics and Computers in Simulation, vol. 55, no. 1, pp. 271 – 280, 2001.

[42] F. Weber and S. Theers, “ODEsensitivity: Sensitivity Analysis of Ordinary Differential Equations,” 2019. R package version 1.1.2.

